# The impact of the two-child benefit cap on parental health in the UK: a triangulation of policy evaluation methods

**DOI:** 10.64898/2026.03.30.26349774

**Authors:** Anabelle Paulino, Jennifer Dykxhoorn, Sara Evans-Lacko, Praveetha Patalay

## Abstract

**Background:** The two-child benefit cap, implemented in April 2017, restricted benefit receipt to the first two children in households with three or more children in the UK. We evaluate the impact of this on parental mental, general, and financial health, as well as investigating how this may differ in particular sociodemographic and economic subgroups.

**Methods:** For this repeated cross-sectional study, we obtained data from 21,741 parents (youngest child aged 5 or under) in the UK Household Longitudinal Survey from 2009 to 2025. Outcomes included parental mental health (psychological distress and life satisfaction), general health (health-related quality of life (HRQoL), self-rated health and health satisfaction), and financial health (financial situation and financial outlook). We used complementary policy evaluation methods to triangulate evidence and strengthen inference: interrupted time series (ITS), difference-in-differences (DiD) and controlled time series analysis (CITS). Subgroup analyses were stratified by sex, ethnicity, and income.

**Results:** Across methods, estimates often lacked precision but the most consistent direction of effects were for parental life satisfaction and financial situation, with some evidence of effects on self-rated health, HRQoL, and financial outlook; and inconsistent estimates for psychological distress and health satisfaction. Impacts were larger among lower-income and ethnic minority parents for some outcomes, while gender differences varied by outcome.

**Conclusions:** Using repeated cross-sectional panel data and triangulating across causal inference methods, we conclude that there is inconsistent and imprecise evidence that the two-child benefit cap in the UK had adverse short-term impacts on health outcomes examined.

## Introduction

Following the 2008 financial crisis, the UK entered a period of spending cuts, including to social benefits [1]. Introduced in April 2017, the two-child benefit cap removed entitlement to Child Tax Credit and Universal Credit for third or subsequent children born on or after 6th April 2017, eliminating what had previously amounted to approximately £3,500 per additional child per year [2,3]. This disproportionately affected families with three or more children, reducing their financial support relative to smaller families and potentially exacerbating financial hardship among some of the most economically vulnerable households in the UK [4].

Research to-date has estimated the two-child benefit cap was associated with a 3 percentage-point increase in maternal employment among affected mothers when their child was aged four, but found no evidence that parents altered birth timing to avoid the policy or that it affected children’s school readiness [5–7]. However, one particularly notable research gap is the impact of this cap on parental health and wellbeing, including mental and financial health. Qualitative research with affected parents has found parents to report adverse experiences including severe financial hardship, increased stress and worry, and the development or worsening of mental health difficulties [8]. Adverse parental mental health has potential implications for child wellbeing and broader societal outcomes including higher health care costs and reduced economic productivity [9–11]. Hence, the primary aim of this study is to estimate the impact of the two-child benefit cap on parental mental, general and financial health outcomes.

The two-child benefit cap has been implicated as a driver of income losses among larger low-income families, with affected households being disproportionately of low income [3,12,13]. For instance, data from the Institute for Fiscal Studies indicates that 91.8% of affected families are in the two lowest income quintiles [3]. Large families from some ethnic minority groups who are more likely to have three or more children are also overrepresented, and qualitative evidence suggests that mothers may experience disproportionate financial strain as a result of the policy [14]. Hence, we will examine whether associations between the benefit cap and parental health differ by sex, ethnicity and family income levels.

In summary, we examine whether the two-child benefit cap had any impact on parental mental, general, or financial health, using UK Household Panel Study data from 2009 to 2025; and whether subgroups of parents based on gender, ethnicity and income were disproportionately impacted. Three complementary quasi-experimental policy evaluation methods, including interrupted time series (ITS) [15], difference-in-differences (DiD) [16], and controlled interrupted time series (CITS) [17] were implemented in order to triangulate inference across methods with different estimands.

## Methods

### Dataset

We used the UK Household Longitudinal Study (UKHLS), which contains information on sociodemographic and economic variables on members of households, including household-level income, and individual health and wellbeing measures [18]. There are up to 15 waves of this repeated-measures cross-sectional survey data, obtained from a combination of yearly face-to-face interviews spanning from 2009 to 2025 [18]. See **Supplement 1** for a timeline mapping years to waves.

The original 2009-2010 sample initially comprised a general population sample from Great Britain of approximately 40,000 households, and in addition to yearly interviews of the original sample has now extended to an additional Ethnic Minority Boost Sample, the former British Household Panel survey, and an Immigrant and Ethnic Minority Boost Sample [19].

Our inclusion criteria were households where at least one resident child was aged 5 or under in each year, and where at least one parent had a response for any of the outcomes. Individuals within the same household, and repeat individuals across multiple years, were treated as separate observations within the dataset. The presence of more than one parent in a household was accounted for in analyses by using the primary sampling unit (PSU) clustering term, which also accounts for household clustering [20].

Based on family size, individuals who had at least one child aged 5 or under were divided into two groups: 3+ and 1-2 child families. The restriction of 5 years of age was chosen to ensure comparability between families with similarly-aged children in both groups, and focus on households most likely to be exposed to the policy’s effects during early childhood, as by five years after policy implementation (2023), all families in the 3+ children group would have been affected by the two-child benefit cap.

Because only children birth year was provided, we excluded individuals whose 3^rd^ child was born in 2017 due to us not being able to ascertain whether these children had been born before or after the benefit cap implementation. **Figure S1** presents the sample flow diagram.

### Intervention

Our intervention was the implementation of the two-child benefit cap on 6^th^ April 2017. Although administered on this specific date, it would take five years for all families in the 3+ children group to have been affected by the exposure as described above due to the intervention cutoff date being based on childbirth after this date. Because the data did not contain enough information to determine individual benefit receipt, our exposure was operationalised as benefit eligibility. To this end, we included a “number of children” variable at each sweep (1-2, or 3+), and derived a variable that specified if individuals were eligible to be affected by the benefit cap (ie 3^rd^ child or later child born after the benefit cap implementation).

We evaluated the impact of the intervention by examining the change in parental outcomes per year in the 3+ children group (ITS), comparing the relative change pre-post intervention in both 3+ and 1-2 children groups (DiD), and comparing the time-trends of parental outcomes per year in each group (CITS) (see **Table 1** for more detail).

**Table 1:**
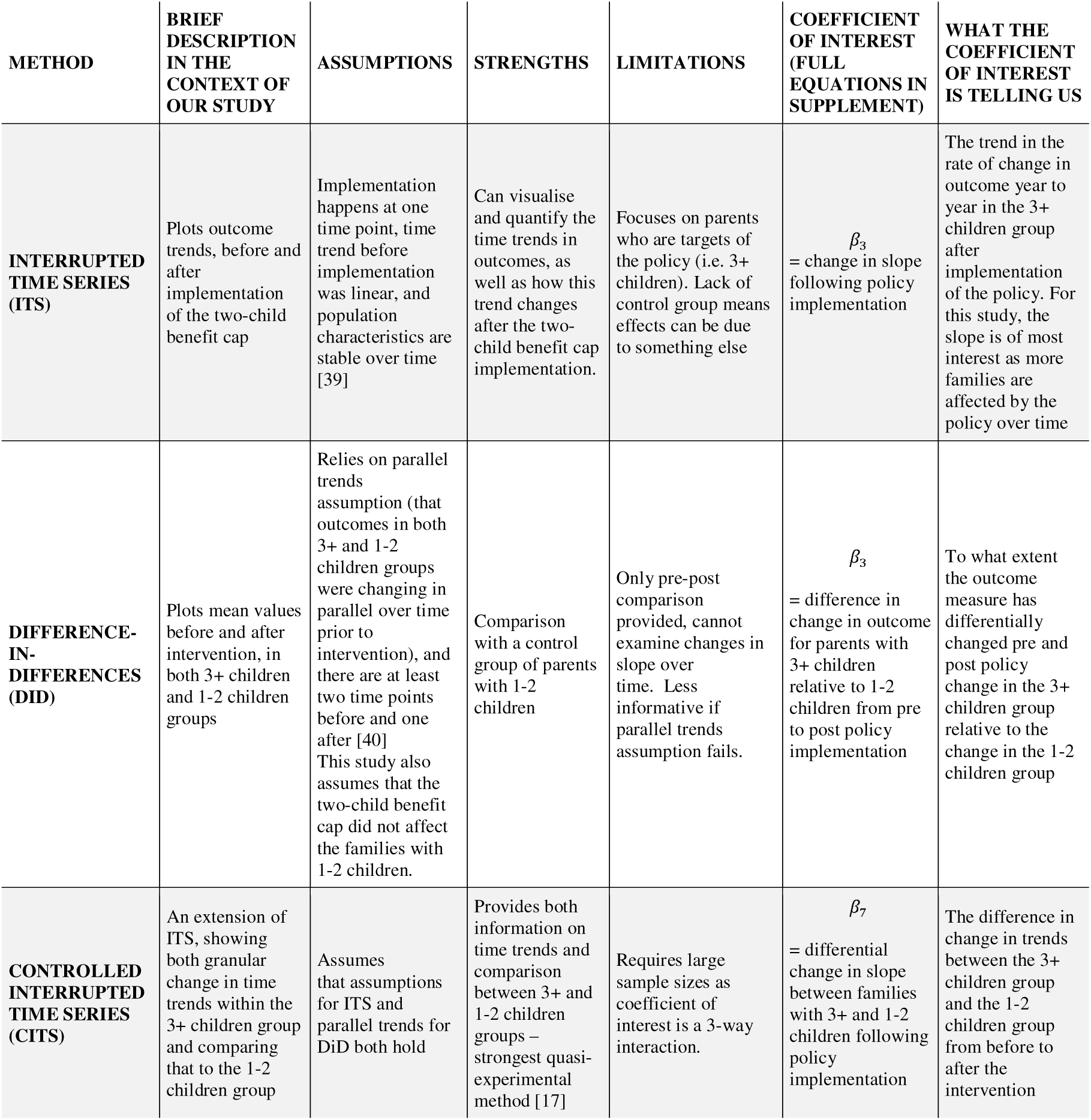
Description of methods used in the study.

### Outcome(s)

We scored all below items in such a way that higher values corresponded to worse outcomes for consistency in directional interpretation of values (see **Supplement 2** for further details of the questions and scoring, and **Figure S2** for correlations).

#### Mental health

For psychological distress, items from the GHQ-12 were summed to create a total continuous score ranging from 0-36, with higher scores indicating greater distress [21].

We evaluated life satisfaction via a single item with possible responses ranging from 1 (“completely satisfied”) to 7 (“completely dissatisfied”).

#### General health

Self-rated health was assessed with a widely-used single item “In general, would you say your health is…”, with responses from 1 (“excellent”) to 5 (“poor”) [22].

Health-related quality of life (HRQoL) was measured using the SF-12, which included questions around physical and mental health, with scores for each questions ranging either from 1-3 or 1-5 based on frequency or severity of limitation [23].

Health satisfaction was measured using a single question, with responses from 1 (“completely satisfied”) to 7 (“completely dissatisfied”).

#### Financial health

Financial situation and financial outlook were each measured with a question, with financial situation having five choices ranging from “living comfortably” to “finding it very difficult” and financial outlook having three responses of “better off”, “about the same”, and “worse off than now”.

### Confounders

There were several confounders included in analyses on the basis of having a potential influence on both family size and parental health. These were sex, country of birth, highest education level, age, ethnicity, marital status, and chronic health conditions. For a more detailed description, see **Supplement 3.**

### Statistical analyses

Power analyses were conducted to determine the minimum detectable effect given the available sample sizes. We used α = 0.1 throughout, as the primary outputs were interaction terms that may be underpowered at the conventional α = 0.05 threshold [24]. Full minimum detectable effects for all outcomes are reported in **Table S1**.

Three complementary analytical approaches were employed (details in **Table 1**, equations in **Supplement 4**). First, interrupted time series (ITS) analysis was conducted in the 3+ children group to examine whether the impact of the two-child benefit cap grew over time as more families became affected. The slope of change in outcomes following policy implementation was the primary measure of interest, and we expected to see an increased slope following the intervention.

Second, difference-in-differences (DiD) analysis was used to estimate the overall policy impact before and after intervention. Prior to DiD analyses, parallel trends testing was conducted per year for main analyses and subgroups. For this DiD, we used a pre-post approach, focusing on 2014-2016 as the “pre” timepoint, as this was the most recent timepoint prior to intervention, and 2023-2025 as the “post” timepoint as all 3+ children families would have been affected by the policy by then.

Because the two-child benefit cap was only implemented for individuals whose 3^rd^ or more child was born after 6^th^ April 2017 (the policy implementation date), individuals whose 3^rd^ or more children were born before the policy implementation would still receive benefits for all their children. We felt that the mixed-exposure affected sample from 2018 to 2022, could lead to a potential dilution of the true before-and-after intercept change in the 3+ children group compared to the 1-2 children control group. In order to address this, we conducted a DiD sensitivity analysis using a propensity-score matched sample [25]. We matched on the number of children in each individual and the age of the youngest child across two time periods (2013-2016 as our “pre” time period and 2018-2023 as our “post” time period). the potential importance of factors such as age of the youngest child and number of children, we felt it informative to create comparable pre-and post-policy groups.

Third, a controlled interrupted time series (CITS) was implemented, incorporating the 1–2 children group as a control primarily to assess consistency in the direction of estimates with the models above. Due to the main estimate of interest being a three-way interaction, we do not expect sufficient precision in effect sizes.

Stratified subgroup analyses were run by sex (male, female), ethnicity (ethnic minority and White), and income quintile (bottom 2 and top 3 quintiles) for both ITS and DiD approaches.

Following UKHLS guidance, population weights were applied in all analyses [26–28]. Because these weights are designed for the UK adult population rather than parents specifically, we ran unweighted sensitivity analyses. As an additional sensitivity analysis, we assessed whether the results reflected a differential COVID-19 impact on larger families^28^ by conducting a COVID-19 DiD analysis using 2019 as the pre-pandemic and 2022 as the post-pandemic time point.

All analyses included unadjusted and adjusted models, with z-score standardisation additionally applied to enable cross-outcome comparisons. Missing data (mainly on confounders) were handled using multiple imputation by chained equations (20 imputations, 15 iterations). Analyses were conducted in R version 4.5.2 [29].

## Results

### Descriptives

There were 76,878 observations (from 21,741 individuals) included in the analytic sample (2009-2025). Out of the total observations across all years included, 57,175 (74.4%) had 1-2 children and 19,703 (25.6%) had 3+ children. See **Table S2** for Ns in each condition at each year among households and individuals. Further descriptives can be found in **Table 2**, with outcome means by family size group and year in **Table S3**.

**Table 2:**
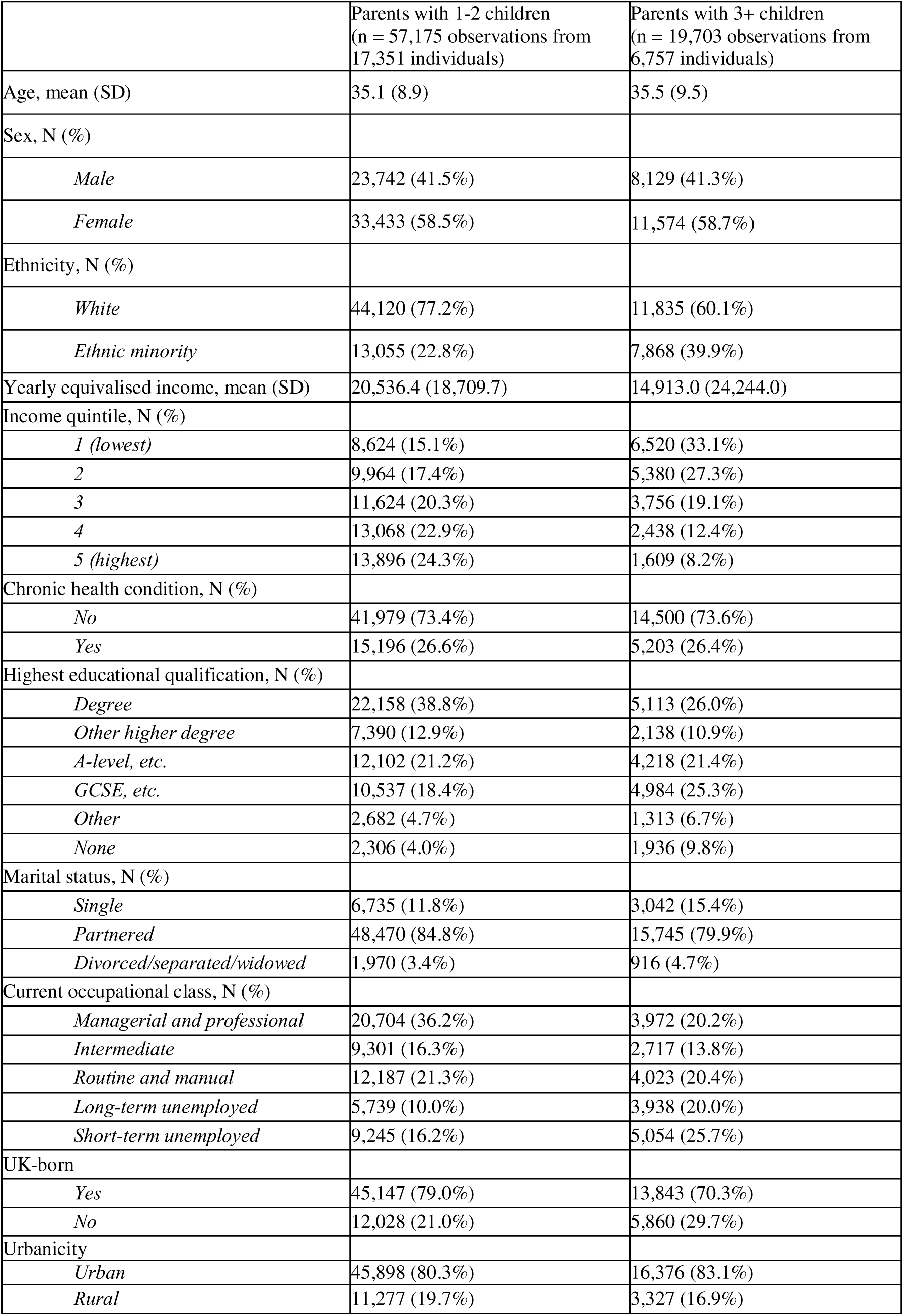
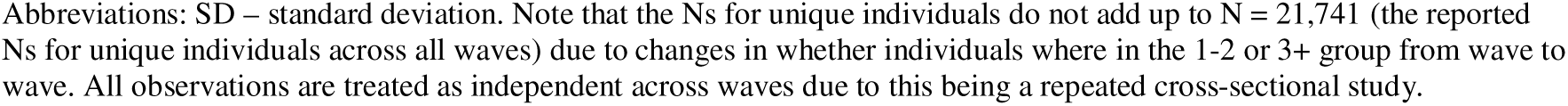
Descriptives by group.

|  | Parents with 1-2 children<br>(n = 57,175 observations from<br>17,351 individuals) | Parents with 3+ children<br>(n = 19,703 observations from<br>6,757 individuals) |
| --- | --- | --- |
| Age, mean (SD) | 35.1 (8.9) | 35.5 (9.5) |
| Sex, N (%) |  |  |
| *Male* | 23,742 (41.5%) | 8,129 (41.3%) |
| *Female* | 33,433 (58.5%) | 11,574 (58.7%) |
| Ethnicity, N (%) |  |  |
| *White* | 44,120 (77.2%) | 11,835 (60.1%) |
| *Ethnic minority* | 13,055 (22.8%) | 7,868 (39.9%) |
| Yearly equivalised income, mean (SD) | 20,536.4 (18,709.7) | 14,913.0 (24,244.0) |
| Income quintile, N (%) |  |  |
| 1 (*lowest*) | 8,624 (15.1%) | 6,520 (33.1%) |
| 2 | 9,964 (17.4%) | 5,380 (27.3%) |
| 3 | 11,624 (20.3%) | 3,756 (19.1%) |
| 4 | 13,068 (22.9%) | 2,438 (12.4%) |
| 5 (*highest*) | 13,896 (24.3%) | 1,609 (8.2%) |
| Chronic health condition, N (%) |  |  |
| *No* | 41,979 (73.4%) | 14,500 (73.6%) |
| *Yes* | 15,196 (26.6%) | 5,203 (26.4%) |
| Highest educational qualification, N (%) |  |  |
| *Degree* | 22,158 (38.8%) | 5,113 (26.0%) |
| *Other higher degree* | 7,390 (12.9%) | 2,138 (10.9%) |
| *A-level, etc.* | 12,102 (21.2%) | 4,218 (21.4%) |
| *GCSE, etc.* | 10,537 (18.4%) | 4,984 (25.3%) |
| *Other* | 2,682 (4.7%) | 1,313 (6.7%) |
| *None* | 2,306 (4.0%) | 1,936 (9.8%) |
| Marital status, N (%) |  |  |
| *Single* | 6,735 (11.8%) | 3,042 (15.4%) |
| *Partnered* | 48,470 (84.8%) | 15,745 (79.9%) |
| *Divorced/separated/widowed* | 1,970 (3.4%) | 916 (4.7%) |
| Current occupational class, N (%) |  |  |
| *Managerial and professional* | 20,704 (36.2%) | 3,972 (20.2%) |
| *Intermediate* | 9,301 (16.3%) | 2,717 (13.8%) |
| *Routine and manual* | 12,187 (21.3%) | 4,023 (20.4%) |
| *Long-term unemployed* | 5,739 (10.0%) | 3,938 (20.0%) |
| *Short-term unemployed* | 9,245 (16.2%) | 5,054 (25.7%) |
| UK-born |  |  |
| *Yes* | 45,147 (79.0%) | 13,843 (70.3%) |
| *No* | 12,028 (21.0%) | 5,860 (29.7%) |
| Urbanicity |  |  |
| *Urban* | 45,898 (80.3%) | 16,376 (83.1%) |
| *Rural* | 11,277 (19.7%) | 3,327 (16.9%) |

Abbreviations: SD – standard deviation. Note that the Ns for unique individuals do not add up to N = 21,741 (the reported Ns for unique individuals across all waves) due to changes in whether individuals were in the 1-2 or 3+ group from wave to wave. All observations are treated as independent across waves due to this being a repeated cross-sectional study.

The proportion of observations with 3+ children affected by the policy in each year from 2018 to 2025 was 23.9% (2018), 42.7% (2019), 56.6% (2020), 76.6% (2021), 91.2% (2022), 100% (2023), 100% (2024), and 100% (2025), see **Figure S3**.

### Are there changes in parental outcomes over time following policy implementation? (Interrupted time series analysis)

Following the introduction of the two-child benefit cap, individuals with 3+ children experienced worsening trends in HRQoL (post-intervention slope increase: 0.08 [90% CI 0.05, 0.11] and financial health (financial situation (0.08 [0.05, 0.11]) and financial outlook (0.04 [0.01, 0.07]) compared with pre-policy trends. Standardised estimates are presented visually in **Figure 1** and numerically in **Table S4**, with raw score estimates in **Figure S4**.

**Figure 1:**
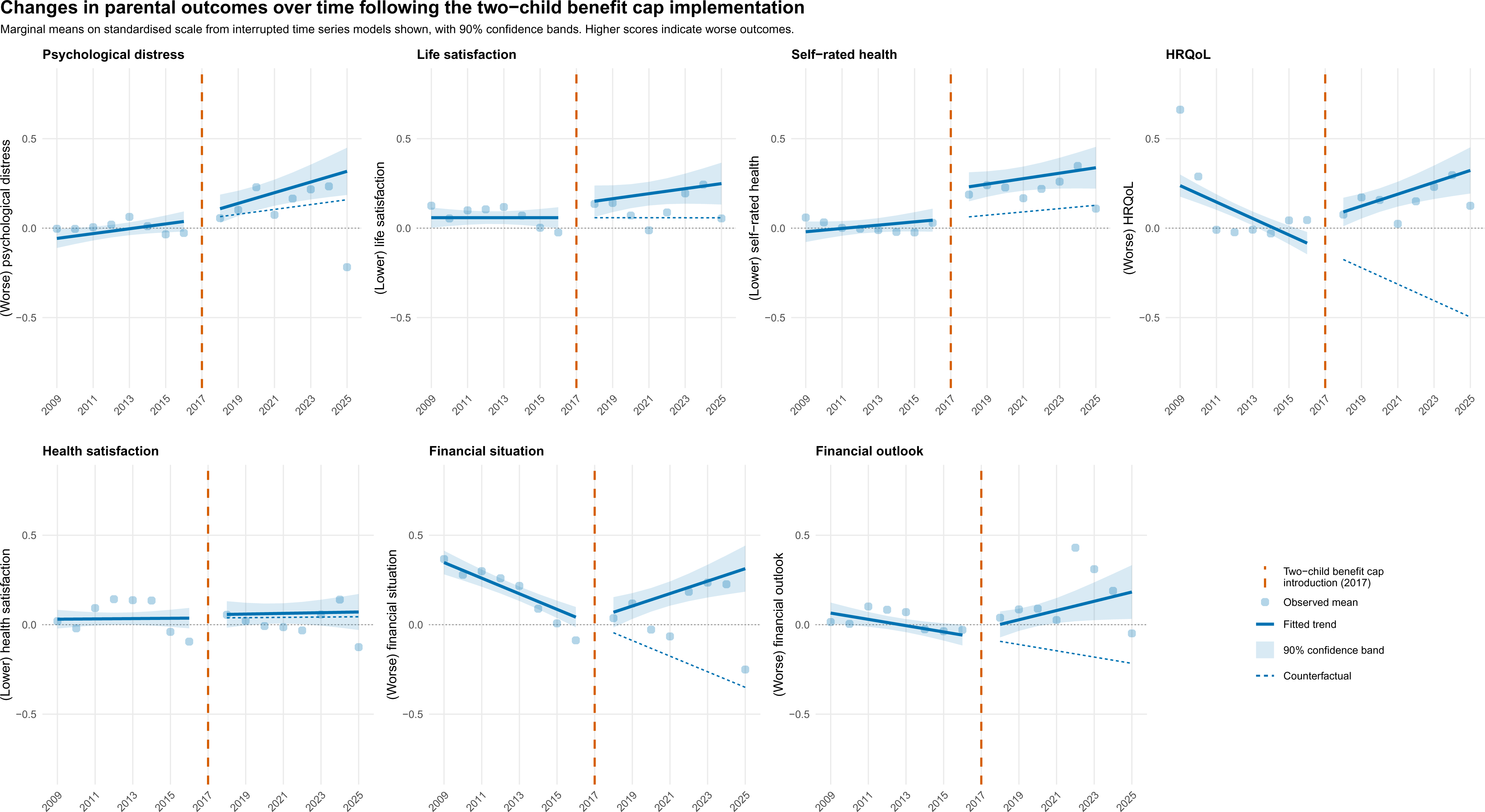
Interrupted time series results across all standardised outcomes with 3+ children group.

Within subgroup analyses, ethnic minority individuals experienced worsening trends for mental health (psychological distress: 0.036 [0.00, 0.07] and life satisfaction: 0.05 [0.01, 0.09]). Individuals in the bottom two income quintiles (0.05 [0.01, 0.09]) and white individuals (0.05 [0.02, 0.08]) showed worsening trends of financial outlook (**Table 3**). Worsening trends of HRQoL and financial situation were seen in all subgroups (**Table 3**).

**Table 3:** ITS (change in slope per year) and DiD (difference in change in outcome) subgroup analyses (90% CIs)

|  |  | Sex |  | Ethnicity |  | Household income |  |
| --- | --- | --- | --- | --- | --- | --- | --- |
|  |  | Male | Female | White | Ethnic minority | Low income | High income |
| Psychological distress | ITS | 0.02 (-0.01, 0.05) | 0.01 (-0.03, 0.05) | 0.01 (-0.02, 0.04) | 0.04 (0.00, 0.07) | 0.02 (-0.01, 0.06) | 0.01 (-0.03, 0.04) |
|  | DiD | -0.01 (-0.20, 0.18) | -0.04 (-0.22, 0.15) | -0.07 (-0.24, 0.11) | 0.09 (-0.15, 0.34) | -0.04 (-0.22, 0.14) | 0.02 (-0.16, 0.20) |
| Life satisfaction | ITS | 0.01 (-0.03, 0.04) | 0.02 (-0.02, 0.05) | 0.00 (-0.03, 0.04) | 0.05 (0.01, 0.09) | 0.03 (-0.01, 0.07) | -0.01 (-0.04, 0.02) |
|  | DiD | 0.07 (-0.18, 0.26) | 0.15 (-0.03, 0.34) | 0.11 (-0.06, 0.28) | 0.19 (-0.07, 0.46) | 0.16 (-0.03, 0.34) | 0.04 (-0.13, 0.20) |
| Self-rated health | ITS | 0.02 (-0.02, 0.05) | -0.00 (-0.04, 0.03) | 0.00 (-0.03, 0.04) | 0.02 (-0.03, 0.07) | 0.02 (-0.02, 0.05) | -0.01 (-0.04, 0.02) |
|  | DiD | 0.08 (-0.09, 0.25) | -0.03 (-0.18, 0.13) | 0.01 (-0.13, 0.14) | 0.08 (-0.14, 0.30) | 0.05 (-0.09, 0.20) | -0.07 (-0.24, 0.09) |
| HRQoL | ITS | 0.09 (0.05, 0.12) | 0.07 (0.03, 0.11) | 0.08 (0.05, 0.12) | 0.08 (0.04, 0.12) | 0.09 (0.06, 0.13) | 0.06 (0.03, 0.09) |
|  | DiD | -0.02 (-0.19, 0.16) | 0.01 (-0.16, 0.18) | 0.00 (-0.17, 0.17) | -0.03 (-0.27, 0.22) | -0.04 (-0.22, 0.14) | 0.01 (-0.16, 0.19) |
| Health satisfaction | ITS | 0.01 (-0.02, 0.04) | -0.01 (-0.04, 0.02) | -0.01 (-0.04, 0.03) | 0.03 (-0.01, 0.06) | 0.00 (-0.03, 0.03) | 0.00 (-0.03, 0.03) |
|  | DiD | -0.01 (-0.15, 0.13) | -0.04 (-0.20, 0.12) | -0.01 (-0.14, 0.12) | 0.01 (-0.21, 0.22) | -0.02 (-0.16, 0.12) | 0.01 (-0.16, 0.19) |
| Financial situation | ITS | 0.06 (0.03, 0.10) | 0.09 (0.06, 0.12) | 0.08 (0.05, 0.11) | 0.08 (0.04, 0.12) | 0.10 (0.06, 0.13) | 0.04 (0.01, 0.08) |
|  | DiD | 0.08 (-0.13, 0.29) | 0.08 (-0.08, 0.25) | 0.14 (-0.04, 0.30) | -0.09 (-0.37, 0.18) | 0.18 (-0.01, 0.37) | -0.03 (-0.25, 0.18) |
| Financial outlook | ITS | 0.05 (0.01, 0.09) | 0.04 (0.01, 0.07) | 0.05 (0.02, 0.08) | 0.03 (-0.02, 0.09) | 0.05 (0.01, 0.09) | 0.03 (-0.00, 0.07) |
|  | DiD | 0.08 (-0.13, 0.29) | 0.04 (-0.11, 0.19) | 0.05 (-0.09, 0.19) | 0.06 (-0.25, 0.36) | 0.09 (-0.10, 0.29) | -0.05 (-0.23, 0.12) |

Abbreviations: ITS – interrupted time series, DiD – difference-in-differences, HRQoL – health related quality of life, CIs – confidence intervals.

### What is the relative change in outcome for parents pre-post policy compared to a control group? (Difference-in-differences analysis)

The parallel trends test between the 2009-2013 and 2014-2016 time periods was numerically passed for all outcomes in main analyses (**Table S5**), although there were some deviations visually (**Figure S5**). Subgroup parallel trends showed deviations for the psychological distress outcome in high-income individuals and for health satisfaction in males and ethnic minority individuals (**Table S6**).

The DiD analysis using a pre (2014-2016) – post (2023-2025) comparison indicated worsening point estimates for the the 3+ children group in life satisfaction (0.12 [-0.02, 0.27]), self-rated health (0.02 [-0.10, 0.14]), and financial health (financial situation (0.09 [-0.06, 0.24]) and financial outlook (0.06 [-0.09, 0.21]) (**Figure 2** (standardised adjusted), **Figure S6** (raw adjusted), **Table S7**), with subgroup analyses showing overall mixed directionality across subgroups and outcomes (**Table 3**).

**Figure 2:**
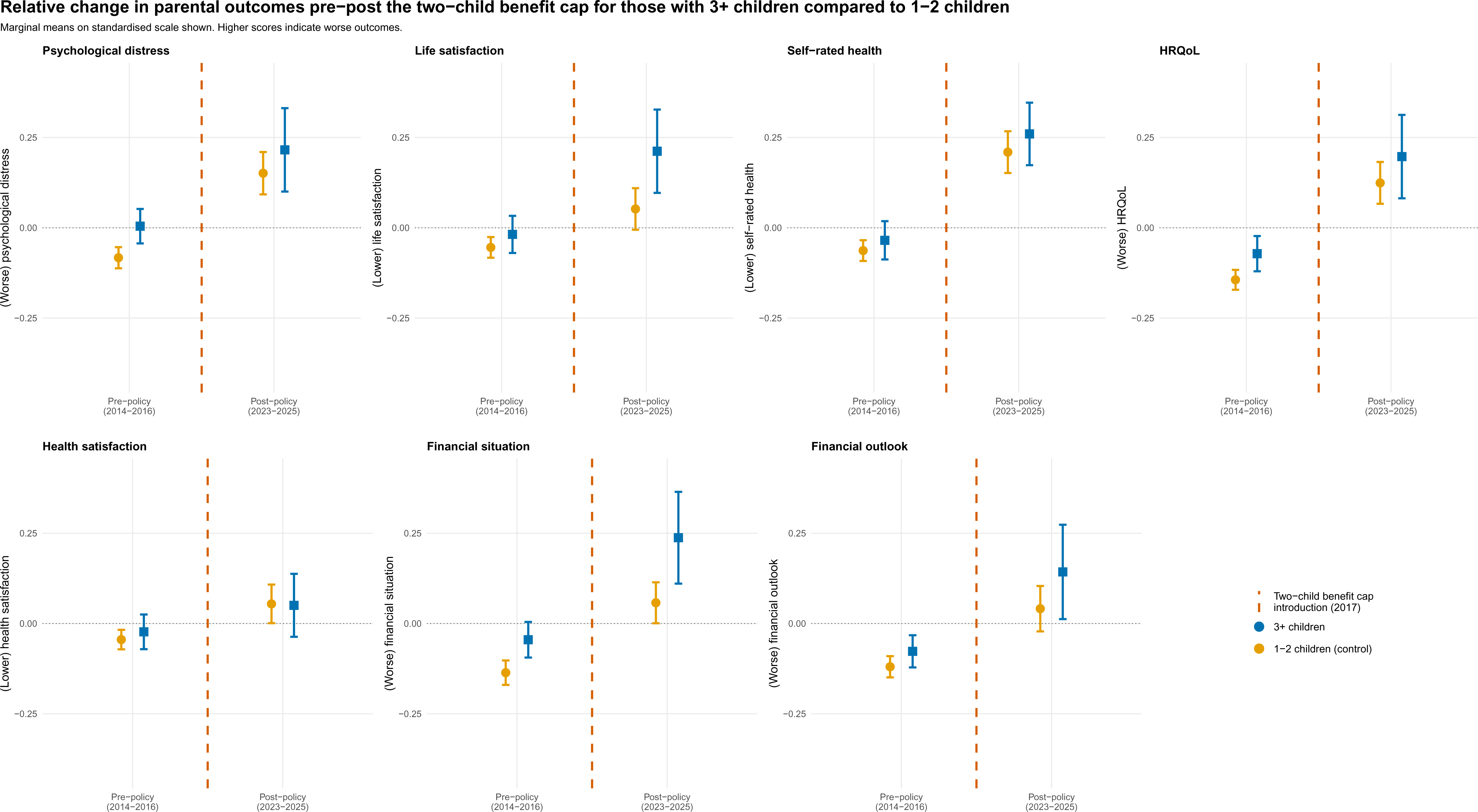
Mean standardised total scores for difference-in-differences analyses between 2014-2016 and 2023-2025.

### Are the changes in the affected families post policy greater than in the control group? (Controlled interrupted time series)

The direction of estimates for the post-policy change in life satisfaction (0.01 [-0.02, 0.04]) and financial situation (0.01 [-0.02, 0.04]) were worsening at a higher rate in the 3+ children group compared to the 1-2 children (**Table S8**, **Figure 3** (standardised adjusted), **Figure S7** (raw adjusted)).

**Figure 3:**
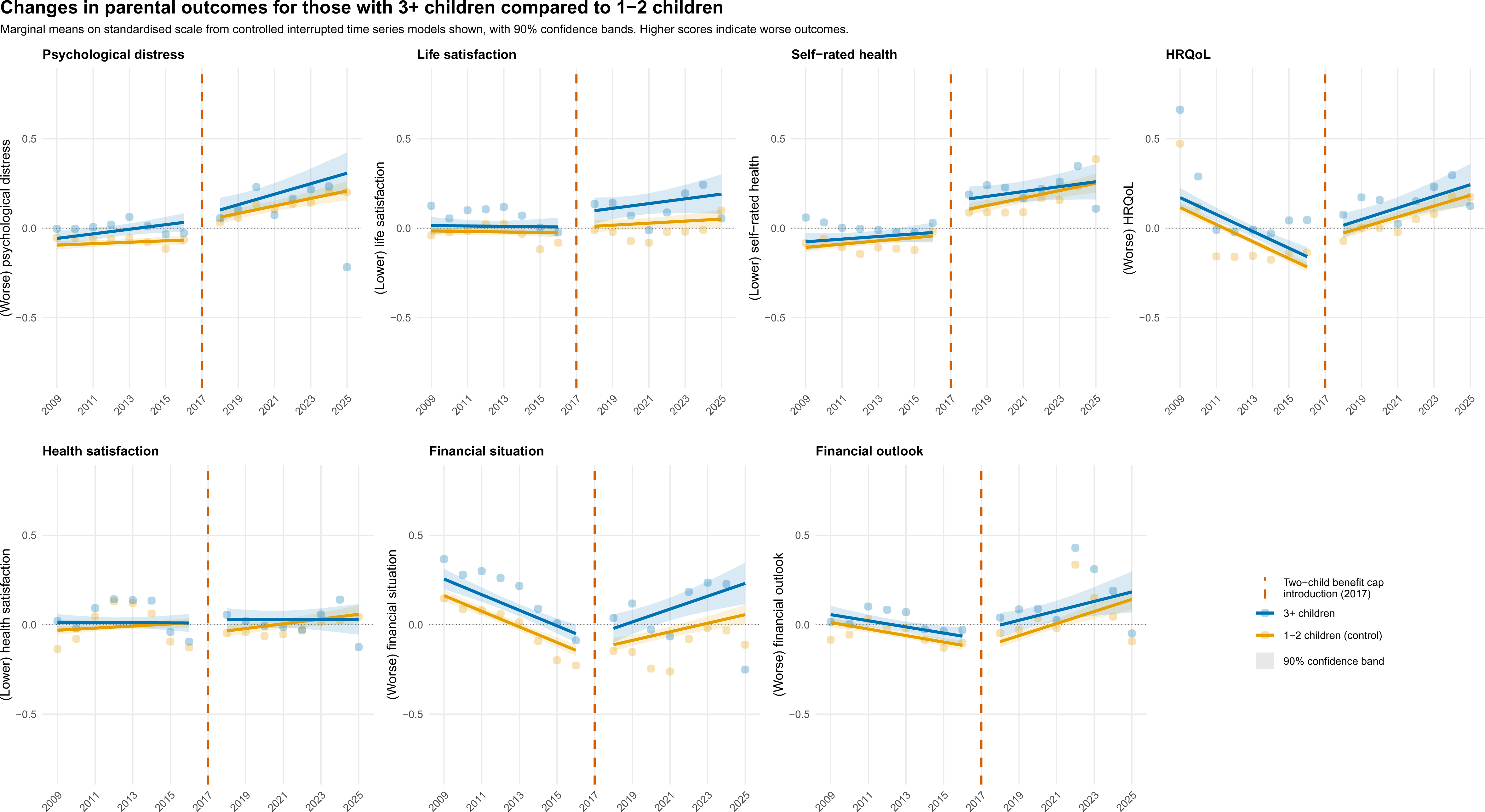
Controlled interrupted time series results across all standardised outcomes with 3+ children compared to 1-2 children group.

### Sensitivity analyses

Results were similar for analyses without panel weights applied, although direction of slope was more consistently worsening in ITS and CITS analyses (**Table S9**), The falsification DiD test showed no measurable differential impact of the COVID-19 pandemic (**Table S10**). The DiD+PSM analysis showed similar results to main DiD analyses (**Table S11**).

### Summary of results

The results triangulated across methods were most directionally consistent for life satisfaction and financial situation, where we find overall worsened direction of estimates for the 3+ families group. Findings were less consistent across methods for psychological distress, self-rated health, and health satisfaction. See **Table 4** for a numerical summary of results for ITS, DiD, and CITS standardised adjusted analyses.

**Table 4:** ITS (change in slope per year), DiD (difference in change in outcome), and CITS (difference in change of slope per year) results for all standardised adjusted outcomes (90% CIs)

|  | ITS | DiD | CITS |
| --- | --- | --- | --- |
|  | Change in slope /<br>year (90% CI) | Family size group indicator<br>* pre-post intervention<br>time indicator (90% CI) | Difference in change in<br>slope of 3+ children<br>group and change in<br>slope of 1-2 children<br>group / year (90% CI) |
| Psychological distress | 0.02 (-0.01, 0.04) | -0.02 (-0.16, 0.12) | -0.00 (-0.03, 0.03) |
| Life satisfaction | 0.01 (-0.01, 0.04) | 0.12 (-0.02, 0.27) | 0.01 (-0.02, 0.04) |
| Self-rated health | 0.01 (-0.02, 0.03) | 0.02 (-0.10, 0.14) | -0.01 (-0.03, 0.02) |
| HRQoL | 0.08 (0.05, 0.11) | 0.00 (-0.14, 0.14) | 0.00 (-0.03, 0.03) |
| Health satisfaction | 0.00 (-0.02, 0.03) | -0.03 (-0.14, 0.09) | -0.01 (-0.03, 0.02) |
| Financial situation | 0.08 (0.05, 0.11) | 0.09 (-0.06, 0.24) | 0.01 (-0.02, 0.04) |
| Financial outlook | 0.04 (0.01, 0.07) | 0.06 (-0.09, 0.21) | -0.01 (-0.04, 0.02) |

Abbreviations: ITS – interrupted time series; DiD – difference in differences; CITS – controlled interrupted time series; CI – confidence interval; HRQoL – health-related quality of life

## Discussion

We used 17 years of UKHLS household panel data (2009-2025) to evaluate the impact of the two-child benefit cap introduced in 2017 on parental mental, general, and financial health using three complementary causal inference methods. Overall, findings were heterogeneous across methods and outcomes. Both life satisfaction and financial situation demonstrated consistently worsening direction of effects post-implementation of the two-child benefit cap across all three methods. The direction and effects were less consistent between methods forfor psychological distress, the general health outcomes, and financial outlook.

The interrupted time series evidence suggests some negative impacts of the policy on parental health outcomes for the 3+ children group. However, these estimates lack precision in analyses including a comparison group (DiD and CITS). Subgroup analyses indicate worsening directionality of several health outcomes for ethnic minorities and lower-income parents, with mixed evidence for differential impacts by gender depending on the outcome.

This policy has been abolished as of April 2026, however, estimating the impact of this policy on a range of outcomes remains important particularly given recent proposals by Reform UK to reimpose the benefit cap should they win the next election [30]. These findings, are aligned with previous evidence of disproportionate impacts of the two child benefit cap based ethnicity and income level on health outcomes [3,13,31]. Our findings on the directional worsening of life satisfaction may also align with evidence from qualitative interviews on the impact of the policy on parental mental health [8]. With the financial situation outcomes being some of the most directionally consistent, this could indicate a potential pathway from financial insecurity to adverse health that warrants further exploration [31,32]. As such, although this study has specifically estimated the immediate policy impacts on parents of young children, there are longer-term consequences on health, wellbeing, and children’s subsequent health that need to be considered.

Strengths of this study included (1) UKHLS, a representative dataset with seventeen years of pre-and post-policy implementation data and rich information on confounders [33,34], (2) comprehensive set of parental health outcomes, and (3) triangulation across complementary causal inference methods, which highlight different parts of the research question and allow for more robust conclusions to be drawn than if any one method had been presented alone.

We were unable to identify potentially policy-exempt parents (e.g. pregnancy resulting from coercion), due to the lack of information on these factors. However, the low proportion of officially-reported exceptions indicates that the proportion of misclassified individuals would likely be low in this study [35]. Due to low power, we were not able to investigate the impact of the two-child benefit cap with regards to specific minoritised ethnic groups, and encourage future research to do so.

Another consideration relating to timing is the COVID-19 pandemic. Given that the policy was introduced 3 years prior to the pandemic, the pandemic’s timing does not interfere with the pre-post policy classification in our study, but could contribute to some of the findings (for instance, increased distress in control group parents in post-policy period). The inclusion of a control group also affected by COVID-19 in the DiD and CITS analyses and results from the COVID-19 DiD test, which estimated differential changes in parental health in the two groups before and after the COVID-19 pandemic, allow us to reasonably conclude that the pandemic does not drive our main findings.

The two-child benefit cap was introduced amongst a wider set of benefits changes, including the overall benefit cap reduction (November 2016), Universal Credit implementation (October 2013), and four-year benefit freeze (April 2016) [36,37]. Given that these policy changes may not overlap in terms of their implementation or target population, we can still reasonably attribute the directionality of impacts in this study to the two-child benefit cap more so than other policies. More broadly, the health impact findings from policy changes to social benefits align with findings from a recent systematic review indicating impacts on mental health of other social benefit changes [38].

Overall, this study shows mixed evidence of worsening mental, general, and financial health trends following the implementation of the two-child benefit cap, with more consistent impacts in ethnic minority and lower-income parents. The magnitude of effects observed in this study should be considered in their population-level context. Even modest changes across a large population could have meaningful implications for health, particularly in the context of wide-reaching social policies. This study underscores the importance of taking into account how policies may affect health, especially when considering its downstream long-term impacts, while also highlighting methodological complexities and benefits of multi-method approaches in estimating the causal impacts of policy changes on health.

## Acknowledgements

We acknowledge the individuals who have participated in the Understanding Society survey. Understanding Society is an initiative funded by the Economic and Social Research Council and various Government Departments, with scientific leadership by the Institute for Social and Economic Research, University of Essex, and survey delivery by the National Centre for Social Research (NatCen) and Verian (formerly Kantar Public). The research data are distributed by the UK Data Service.

## Statements and Declarations

### Author contributions

Conceptualisation: AP, PP

Methodology: AP, PP, JD, SEL

Formal analysis: AP

Visualisation: AP

Writing-Original draft: AP, PP

Writing-Revision: AP, PP, JD, SEL

Supervision: PP, JD, SEL

All authors read and approved the final manuscript.

### Ethics approval

We have used data from the Understanding Society survey, which has been approved by the University of Essex Ethics Committee. Please see the following link for ethics approval and informed consent information: https://www.understandingsociety.ac.uk/documentation/mainstage/user-guides/main-survey-user-guide/ethics/

### Data availability

Data is available for download from UK Data Service following registration at the following link: https://datacatalogue.ukdataservice.ac.uk/studies/study/6614#details.

## References

1. Emmerson C, Johnson P, Ridpath N. The Conservatives and the Economy, 2010–24. Institute for Fiscal Studies; 2024 [cited 2025 Oct 29]; 10.1920/wp.ifs.2024.2424

2. Department for Work and Pensions, HM Revenue and Customs. Families with more than 2 children: claiming benefits [Internet]. GOV.UK. 2017 [cited 2026 Feb 20]. https://www.gov.uk/guidance/claiming-benefits-for-2-or-more-children. Accessed 20 Feb 2026

3. Waters T, Latimer E. The two-child limit: poverty, incentives and cost [Internet]. Inst. Fisc. Stud. 2024 [cited 2025 Oct 29]. https://ifs.org.uk/articles/two-child-limit-poverty-incentives-and-cost. Accessed 29 Oct 2025

4. Department for Work & Pensions. Removing the two-child limit on Universal Credit: Impact on low income poverty levels in the United Kingdom [Internet]. GOV.UK. 2025 [cited 2026 Feb 20]. https://www.gov.uk/government/publications/poverty-impacts-of-social-security-changes-at-budget-2025/removing-the-two-child-limit-on-universal-credit-impact-on-low-income-poverty-levels-in-the-united-kingdom. Accessed 20 Feb 2026

5. Cook W. Unconditional Cash Transfers and Maternal Labour Supply: The Case of the Two Child Limit in the UK [Internet]. 2025 [cited 2025 Oct 29]. 10.2139/ssrn.5163890

6. Reader M, Portes J, Patrick R. Does Cutting Child Benefits Reduce Fertility in Larger Families? Evidence from the UK’s Two-Child Limit. Popul Res Policy Rev. 2025;44:21. 10.1007/s11113-025-09935-5

7. Cattan S, Wernham T, Waters T. What is the effect of the two-child limit on children’s school readiness? [Internet]. Institute for Fiscal Studies; 2025 [cited 2025 Oct 29]. 10.1920/re.ifs.2025.0028

8. Motiani S, Lucas PJ, Bazalgette L. Lost opportunities: parents’ perspectives on how the two-child limit policy is affecting their children’s early learning and development [Internet]. Nesta; 2024 Apr. https://media.nesta.org.uk/documents/Lost_opportunities_-_parents_perspectives_on_how_the_two-child_limit_policy_is_affecting_their_childrens_early_learning_and_development_JiZp7LL.pdf

9. Kamis C. The Long-Term Impact of Parental Mental Health on Children’s Distress Trajectories in Adulthood. Soc Ment Health. SAGE Publications Inc; 2021;11:54–68. 10.1177/2156869320912520

10. Brown CC, Adams CE, George KE, Moore JE. Mental Health Conditions Increase Severe Maternal Morbidity By 50 Percent And Cost $102 Million Yearly In The United States. Health Aff Proj Hope. 2021;40:1575–84. 10.1377/hlthaff.2021.00759

11. Luca DL, Margiotta C, Staatz C, Garlow E, Christensen A, Zivin K. Financial Toll of Untreated Perinatal Mood and Anxiety Disorders Among 2017 Births in the United States. Am J Public Health. 2020;110:888–96. 10.2105/AJPH.2020.305619

12. Mansell E. The two-child limit to means tested benefits [Internet]. Womens Budg. Group. 2023 [cited 2026 Feb 20]. https://www.wbg.org.uk/publication/the-two-child-limit-to-means-tested-benefits/. Accessed 20 Feb 2026

13. Chzhen Y, Bradshaw J. The two-child limit and child poverty in the United Kingdom. Int J Soc Welf. 2025;34:e12642. 10.1111/ijsw.12642

14. Andersen K. Promoting fairness? Exploring the gendered impacts of the benefit cap and the two-child limit. J Poverty Soc Justice. Policy Press; 2023;31:174–90. 10.1332/175982721X16757603309669

15. Bernal JL, Cummins S, Gasparrini A. Interrupted time series regression for the evaluation of public health interventions: a tutorial. Int J Epidemiol. 2017;46:348–55. 10.1093/ije/dyw098

16. Rothbard S, Etheridge JC, Murray EJ. A Tutorial on Applying the Difference-in-Differences Method to Health Data. Curr Epidemiol Rep. 2023;11:85–95. 10.1007/s40471-023-00327-x

17. Lopez Bernal J, Cummins S, Gasparrini A. The use of controls in interrupted time series studies of public health interventions. Int J Epidemiol. 2018;47:2082–93. 10.1093/ije/dyy135

18. University of Essex I for S and ER. Understanding Society: Waves 1-14, 2009-2023 and Harmonised BHPS: Waves 1-18, 1991-2009 [Internet]. UK Data Service; 2024 [cited 2026 Feb 10]. 10.5255/UKDA-SN-6614-20

19. University of Essex. Main survey [Internet]. Underst. Soc. [cited 2026 Feb 20]. https://www.understandingsociety.ac.uk/documentation/mainstage/. Accessed 20 Feb 2026

20. National Centre for Research Methods. Clustering and PSU’s (Primary Sampling Units) [Internet]. 2022 [cited 2026 Aug 28]. https://www.restore.ac.uk/PEAS/clustering.php. Accessed 28 Aug 2026

21. Goldberg DP, Williams P. A User’s Guide to the General Health Questionnaire. NFER-NELSON; 1988.

22. Idler EL, Benyamini Y. Self-rated health and mortality: a review of twenty-seven community studies. J Health Soc Behav. 1997;38:21–37.

23. Ware JE, Kosinski M, Keller SD. A 12-Item Short-Form Health Survey: Construction of Scales and Preliminary Tests of Reliability and Validity. Med Care. 1996;34:220.

24. Marshall SW. Power for tests of interaction: effect of raising the Type I error rate. Epidemiol Perspect Innov. 2007;4:4. 10.1186/1742-5573-4-4

25. Benedetto U, Head SJ, Angelini GD, Blackstone EH. Statistical primer: propensity score matching and its alternatives. Eur J Cardiothorac Surg. 2018;53:1112–7. 10.1093/ejcts/ezy167

26. University of Essex. Longitudinal and cross-sectional weights [Internet]. Underst. Soc. [cited 2026 Aug 31]. https://www.understandingsociety.ac.uk/documentation/mainstage/user-guides/main-survey-user-guide/longitudinal-and-cross-sectional-weights/. Accessed 31 Aug 2026

27. University of Essex. Weighting guidance [Internet]. Underst. Soc. [cited 2026 Aug 31]. https://www.understandingsociety.ac.uk/documentation/mainstage/user-guides/main-survey-user-guide/weighting-guidance/. Accessed 31 Aug 2026

28. University of Essex. Selecting the correct weight for your analysis [Internet]. Underst. Soc. [cited 2026 Aug 31]. https://www.understandingsociety.ac.uk/documentation/mainstage/user-guides/main-survey-user-guide/selecting-the-correct-weight-for-your-analysis/. Accessed 31 Aug 2026

29. R Core Team. R: The R Project for Statistical Computing [Internet]. Vienna, Austria: R Foundation for Statistical Computing; 2025 [cited 2026 Feb 20]. https://www.r-project.org/. Accessed 20 Feb 2026

30. Nevett J. Reform UK would reimpose two-child benefit cap [Internet]. BBC News. 2026 [cited 2026 Feb 20]. https://www.bbc.co.uk/news/articles/c5yrgp0pjlvo. Accessed 20 Feb 2026

31 . Marmot M. Health equity in England: the Marmot review 10 years on. BMJ. British Medical Journal Publishing Group; 2020;368:m693. 10.1136/bmj.m693

32. Thomson RM, Igelström E, Purba AK, Shimonovich M, Thomson H, McCartney G, et al. How do income changes impact on mental health and wellbeing for working-age adults? A systematic review and meta-analysis. Lancet Public Health. 2022;7:e515–28. 10.1016/S2468-2667(22)00058-5

33. Platt L, Knies G, Luthra R, Nandi A, Benzeval M. Understanding Society at 10 Years. Eur Sociol Rev. 2020;36:976–88. 10.1093/esr/jcaa031

34. Buck N, McFall S. Understanding Society: design overview. Longitud Life Course Stud. 2012;3:5–17. 10.14301/llcs.v3i1.159

35. Department for Work and Pensions. Universal Credit claimants statistics on the two child limit policy, April 2025 [Internet]. GOV.UK. 2025 [cited 2026 Feb 20]. https://www.gov.uk/government/statistics/universal-credit-claimants-statistics-on-the-two-child-limit-policy-april-2025. Accessed 20 Feb 2026

36. Beasor S. Welfare benefits: the reform timeline [Internet]. HQN; 2026. https://hqnetwork.co.uk/wp-content/uploads/2026/02/Welfare-reform-timeline-2026.pdf

37. HM Treasurer, The Rt Hon George Osborne. Chancellor George Osborne’s Summer Budget 2015 speech [Internet]. GOV.UK. 2015 [cited 2025 Oct 29]. https://www.gov.uk/government/speeches/chancellor-george-osbornes-summer-budget-2015-speech. Accessed 29 Oct 2025

38. Simpson J, Albani V, Bell Z, Bambra C, Brown H. Effects of social security policy reforms on mental health and inequalities: A systematic review of observational studies in high-income countries. Soc Sci Med. 2021;272:113717. 10.1016/j.socscimed.2021.113717

39. Kontopantelis E, Doran T, Springate DA, Buchan I, Reeves D. Regression based quasi-experimental approach when randomisation is not an option: interrupted time series analysis. The BMJ. 2015;350:h2750. 10.1136/bmj.h2750

40. Schwerdt G, Woessmann L. Chapter 1 - Empirical methods in the economics of education. In: Bradley S, Green C, editors. Econ Educ Second Ed [Internet]. Academic Press; 2020 [cited 2025 Oct 29]. p. 3–20. 10.1016/B978-0-12-815391-8.00001-X

