## Supplement for "The impact of the two-child benefit cap on parental health in the UK: a triangulation of policy evaluation methods"

### Supplement 1: Timeline of waves in each year

The timeline below is adapted from the UK Household Longitudinal Study (UKHLS) website.^1^

|  | 2009 | 2010 | 2011 | 2012 | 2013 | 2014 | 2015 | 2016 | 2017 | 2018 | 2019 | 2020 | 2021 | 2022 | 2023 | 2024 | 2025 |
| --- | --- | --- | --- | --- | --- | --- | --- | --- | --- | --- | --- | --- | --- | --- | --- | --- | --- |
| Wave 1 |  |  |  |  |  |  |  |  |  |  |  |  |  |  |  |  |  |
| Wave 2 |  |  |  |  |  |  |  |  |  |  |  |  |  |  |  |  |  |
| Wave 3 |  |  |  |  |  |  |  |  |  |  |  |  |  |  |  |  |  |
| Wave 4 |  |  |  |  |  |  |  |  |  |  |  |  |  |  |  |  |  |
| Wave 5 |  |  |  |  |  |  |  |  |  |  |  |  |  |  |  |  |  |
| Wave 6 |  |  |  |  |  |  |  |  |  |  |  |  |  |  |  |  |  |
| Wave 7 |  |  |  |  |  |  |  |  |  |  |  |  |  |  |  |  |  |
| Wave 8 |  |  |  |  |  |  |  |  |  |  |  |  |  |  |  |  |  |
| Wave 9 |  |  |  |  |  |  |  |  |  |  |  |  |  |  |  |  |  |
| Wave 10 |  |  |  |  |  |  |  |  |  |  |  |  |  |  |  |  |  |
| Wave 11 |  |  |  |  |  |  |  |  |  |  |  |  |  |  |  |  |  |
| Wave 12 |  |  |  |  |  |  |  |  |  |  |  |  |  |  |  |  |  |
| Wave 13 |  |  |  |  |  |  |  |  |  |  |  |  |  |  |  |  |  |
| Wave 14 |  |  |  |  |  |  |  |  |  |  |  |  |  |  |  |  |  |
| Wave 15 |  |  |  |  |  |  |  |  |  |  |  |  |  |  |  |  |  |

### Supplement 2: Questionnaires from Understanding Society dataset

This version contains questions in the order they were scored for analyses (in other words, reverse-scored as necessary).

#### General Health Questionnaire (GHQ-12)

A) GHQ: concentration

The next questions are about how you have been feeling over the last few weeks. Have you recently been able to concentrate on whatever you're doing?

1 Better than usual
2 Same as usual
3 Less than usual
4 Much less than usual
B) GHQ: loss of sleep

Have you recently lost much sleep over worry?

1 Not at all
2 No more than usual
3 Rather more than usual
4 Much more than usual
C) GHQ: playing a useful role

Have you recently felt that you were playing a useful part in things?

1 More so than usual
2 Same as usual
3 Less so than usual
4 Much less than usual
D) GHQ: capable of making decisions

Have you recently felt capable of making decisions about things?

1 More so than usual
2 Same as usual
3 Less so than usual
4 Much less than usual
E) GHQ: constantly under strain

Have you recently felt constantly under strain?

1 Not at all
2 No more than usual
3 Rather more than usual
4 Much more than usual
F) GHQ: problem overcoming difficulties

Have you recently felt you couldn’t overcome your difficulties?

1 Not at all
2 No more than usual
3 Rather more than usual
4 Much more than usual
G) GHQ: enjoy day-to-day activities

Have you recently been able to enjoy your normal day-to-day activities?

1 More so than usual
2 Same as usual
3 Less so than usual
4 Much less than usual
H) GHQ: ability to face problems

Have you recently been able to face up to problems?

1 More so than usual
2 Same as usual
3 Less so than usual
4 Much less than usual
I) GHQ: unhappy and depressed

Have you recently been feeling unhappy or depressed?

1 Not at all
2 No more than usual
3 Rather more than usual
4 Much more than usual
J) GHQ: losing confidence

Have you recently been losing confidence in yourself?

1 Not at all
2 No more than usual
3 Rather more than usual
4 Much more than usual
K) GHQ: believe worthless

Have you recently been thinking of yourself as a worthless person?

1 Not at all
2 No more than usual
3 Rather more than usual
4 Much more than usual
L) GHQ: general happiness

Have you recently been feeling reasonably happy, all things considered?

1 More so than usual
2 Same as usual
3 Less so than usual
4 Much less than usual

#### Short Form (SF-12) Questionnaire

1) General health

In general, would you say your health is…

1 Excellent
2 Very good
3 Good
4 Fair
5 Poor

2a) Health limits moderate activities

The following questions are about activities you might do during a typical day. Does your health now limit you in these activities? If so, how much?

Moderate activities, such as moving a table, pushing a vacuum cleaner, bowling, or playing golf

1 No, not limited at all
2 Yes, limited a little
3 Yes, limited a lot

2b) Health limits several flights of stairs

Climbing several flights of stairs

1 No, not limited at all
2 Yes, limited a little
3 Yes, limited a lot

3a) Physical health limits amount of work

During the past 4 weeks, how much of the time have you had any of the following problems with your work or other regular daily activities as a result of your physical health?

Accomplished less than you would like

1 None of the time
2 A little of the time
3 Some of the time
4 Most of the time
5 All of the time

3b) Physical health limits kind of work

Were limited in the kind of work or other activities

1 None of the time
2 A little of the time
3 Some of the time
4 Most of the time
5 All of the time

4a) Last 4 weeks: Mental health meant accomplished less

During the past 4 weeks, how much of the time have you had any of the following problems with your work or other regular daily activities as a result of any emotional problems (such as feeling depressed or anxious)?

Accomplished less than you would like

1 None of the time
2 A little of the time
3 Some of the time
4 Most of the time
5 All of the time

4b) Last 4 weeks: Mental health meant worked less carefully

Did work or other activities less carefully than usual

1 None of the time
2 A little of the time
3 Some of the time
4 Most of the time
5 All of the time

5) Last 4 weeks: Pain interfered with work

During the past 4 weeks, how much did pain interfere with your normal work (including both work outside the home and housework)?

1 Not at all
2 A little bit
3 Moderately
4 Quite a bit
5 Extremely

6a) Last 4 weeks: Felt calm and peaceful

These questions are about how you feel and how things have been with you during the past 4 weeks. For each question, please give the one answer that comes closest to the way you have been feeling. How much of the time during the past 4 weeks...

Have you felt calm and peaceful?

1 All of the time
2 Most of the time
3 Some of the time
4 A little of the time
5 None of the time

6b) Last 4 weeks: Had a lot of energy

Did you have a lot of energy?

1 All of the time
2 Most of the time
3 Some of the time
4 A little of the time
5 None of the time

6c) Last 4 weeks: Felt downhearted and depressed

Have you felt downhearted and depressed?

1 None of the time
2 A little of the time
3 Some of the time
4 Most of the time
5 All of the time

7) Last 4 weeks: Physical or mental health interfered with social life

During the past 4 weeks, how much of the time has your physical health or emotional problems interfered with your social activities (like visiting friends, relatives)?

1 None of the time
2 A little of the time
3 Some of the time
4 Most of the time
5 All of the time

#### Satisfaction with health

Here are some questions about how you feel about your life. Please choose the number which you feel best describes how dissatisfied or satisfied you are with the following aspects of your current situation.

Your health.

1 Completely satisfied
2 Mostly satisfied
3 Somewhat satisfied
4 Neither Sat not Dissat
5 Somewhat dissatisfied
6 Mostly dissatisfied
7 Completely dissatisfied

#### Satisfaction with life overall

Your life overall.

1 Completely satisfied
2 Mostly satisfied
3 Somewhat satisfied
4 Neither Sat not Dissat
5 Somewhat dissatisfied
6 Mostly dissatisfied
7 Completely dissatisfied

#### Subjective financial situation

How well would you say you yourself are managing financially these days?

Would you say you are…

1 Living comfortably
2 Doing alright
3 Just about getting by
4 Finding it quite difficult
5 Finding it very difficult

#### Subjective financial outlook

Looking ahead, how do you think you will be financially a year from now, will you be…

1 Better off
2 About the same
3 Worse off than now

### Supplement 3: Description of confounders used in study

#### Time-invariant confounders

Confounders which are time-invarying included country of birth (with a binary variable indicating whether individuals were born in the UK or not), highest education level (using a derived variable with standard groupings of “Degree”, “Other higher degree”, “A-level, etc.”, “GSCE, etc”, “Other” or “None”), religious affiliation, ethnicity (for power reasons, classified as White (defined by indicating either “British/English/Scottish/Welsh/Northern Irish”, “Irish”, or “Any other White background”) versus any other ethnicity), and sex (with a binary variable indicating “male” or “female”). These data were collected at the first available sweep for each included individual.

#### Time-variant confounders

Confounders that are time-varying from each wave included age (continuous), urbanicity (a binary derived variable, with “1” indicating that the address is located within an urban settlement of 10,000 or more people), de facto marital status (a derived variable grouped into “Single”, “Partnered”, and “Divorced/separated/widowed” categorisations), the presence of a current or past chronic health condition, and whether individuals included had ever been diagnosed with a disability.

We decided to exclude the confounders of religious affiliation and disability status at the imputation on the basis of having >30% missingness.

We display information on occupation class (a derived variable with NS-SEC coding, grouped into “Managerial and professional”, “Intermediate”, “Routine and manual”, “Long-term unemployed” (defined as 3+ waves of responding “unemployed”), and “Short-term unemployed” (defined as 1-2 waves of responding “unemployed”)) and household income (an equivalised value calculated by dividing each total household income value by an OECD-modified equivalence scale for each household), but have not included those terms in analyses due to them being on the causal pathway between the two-child benefit cap implementation and parental mental, general, or financial health.

### Supplement 4: Equations for the different models

For each equation, the coefficient of interest is listed in bold.

#### Interrupted time series (ITS)

$$Y_{i,h, t}= \beta_{0}+ \beta_{1}X_{1t}+ \beta_{2}X_{2t}+ \beta_{3}\left( X_{1t} \cdot X_{2t} \right)+confounders (in adjusted models only)\mathbf{+}\in_{i,h,t}$$

Where:

$Y_{i,h,t}$ = outcome for individual *i* in household *h* at time point *t*

$\beta_{0}$ = intercept

$\beta_{1}$ = pre-policy time trend

$X_{1t}$ = time since policy (in years, with 0 being April 2017)

$\beta_{2}$ = change at policy implementation

$X_{2t}$ = pre or post policy indicator (with 0= pre and 1 = post April 2017)

$\boldsymbol{\beta}_{\boldsymbol{3}}$**= change in slope following policy implementation**

$X_{1t} \cdot X_{2t}$ = time since policy * pre/post policy indicators

$\in_{i,h,t}$ = clustered household error term

#### Difference-in-differences (DiD)

$$Y_{i,h,t}= \beta_{0}+ \beta_{1}X_{1}+ \beta_{2}X_{2t}+ \beta_{3}\left( X_{1} \cdot X_{2t} \right)+confounders (in adjusted models only)\mathbf{+}\in_{i,h, t}$$

Where:

$Y_{i,h,t}$ = outcome for individual *i* in household *h* at time point *t*

$\beta_{0}$ = intercept

$\beta_{1}$ = baseline difference in outcome between 3+ and 1-2 children groups

$X_{1}$ = family size indicator (1/0, with 1 = 3+ children and 0 = 1-2 children)

$\beta_{2}$ = change in outcome common to both groups

$X_{2t}$ = time period indicator (1/0, with 1 = “pre” time period and 0 = “post” time period)

$\boldsymbol{\beta}_{\boldsymbol{3}}$**= difference in change in outcome for parents with 3+ children relative to 1-2 children from pre to post policy implementation**

$\in_{i,h,t}$ = clustered household error term

#### Controlled interrupted time series (CITS)

$$Y_{i,h,t}= \beta_{0}+ \beta_{1}X_{1t}+ \beta_{2}X_{2t}+\beta_{3}X_{3}+ \beta_{4}\left( X_{1t} \cdot X_{2t} \right)+\beta_{5}\left( X_{1t} \cdot X_{3} \right)+\beta_{6}\left( X_{2t} \cdot X_{3} \right)+ \beta_{7}\left( {X_{1t}\cdot X}_{2t} \cdot X_{3} \right)+confounders (in adjusted models only)\mathbf{+}\in_{i,h, t}$$

$Y_{i,h,t}$ = outcome for individual *i* in household *h* at time point *t*

$\beta_{0}$ = intercept

$\beta_{1}$ = pre-policy time trend

$X_{1t}$ = time since policy (in years, with 0 being April 2017)

$\beta_{2}$ = change at policy implementation

$X_{2t}$ = pre or post policy indicator ( with 0= pre and 1 = post April 2017)

$\beta_{3}$ = baseline difference in outcome between 3+ and 1-2 children groups

$X_{3}$ = family size indicator (1/0, with 1 = 3+ children and 0 = 1-2 children)

$\beta_{4}$= change in slope following policy implementation common to both groups

$X_{1t} \cdot X_{2t}$ = time since policy * pre/post policy indicator

$\beta_{5}$= difference in pre-post policy time trends between 3+ and 1-2 children groups

$X_{1t} \cdot X_{3}$ = time since policy * family size indicator

$\beta_{6}$= difference in change in outcome between 3+ and 1-2 children groups following policy implementation

$X_{2t} \cdot X_{3}$ = pre/post policy * family size indicators

$\boldsymbol{\beta}_{\boldsymbol{7}}$**= differential change in slope between families with 3+ and 1-2 children following policy implementation**

$X_{1t} \cdot X_{2t}\cdot X_{3}$ = time since policy * pre-post policy * family size indicators

$\in_{i,h,t}$ = clustered household error term

### Supplementary Tables

#### Table S1: Minimum detectable effects

Due to the clustered nature of analyses, we first present the intraclass correlation coefficient (ICC) of each outcome. We then calculate the design effect size, which varies by method and is a combination of ICC and the average cluster size (in this case, the average number of observations in each primary sampling unit (PSU)). The power analysis results are then adjusted by each design effect size to result in the minimum detectable effects (MDEs) presented here. The MDEs are presented as changes in standard deviations to reflect the standardised nature of the analyses across outcomes and methods.

| Outcome | ICC | Interrupted Time Series (ITS) MDE | Difference-in-Differences (DiD) MDE | Controlled Interrupted Time Series (CITS) MDE | Falsification DiD (sensitivity) MDE | DiD with propensity score matching (sensitivity) MDE |
| --- | --- | --- | --- | --- | --- | --- |
| Psychological distress | 0.148 | 0.026 | 0.134 | 0.036 | 0.185 | 0.092 |
| Life satisfaction | 0.143 | 0.026 | 0.133 | 0.036 | 0.185 | 0.091 |
| Self-rated health | 0.193 | 0.028 | 0.142 | 0.040 | 0.192 | 0.100 |
| HRQoL | 0.200 | 0.029 | 0.143 | 0.040 | 0.193 | 0.101 |
| Health satisfaction | 0.112 | 0.025 | 0.127 | 0.033 | 0.180 | 0.086 |
| Financial situation | 0.255 | 0.031 | 0.153 | 0.044 | 0.201 | 0.110 |
| Financial outlook | 0.113 | 0.025 | 0.128 | 0.033 | 0.180 | 0.086 |

#### Table S2: Sample size per year

| Year |  | 1-2 children | 3+ children (unaffected by policy) | 3+ children (affected by policy) |
| --- | --- | --- | --- | --- |
| 2009 | Households | 1876 | 615 | - |
|  | Observations | 3040 | 1056 | - |
| 2010 | Households | 3729 | 1353 | - |
|  | Observations | 6124 | 2388 | - |
| 2011 | Households | 3379 | 1272 | - |
|  | Observations | 5641 | 2271 | - |
| 2012 | Households | 2993 | 1099 | - |
|  | Observations | 5095 | 1927 | - |
| 2013 | Households | 2631 | 973 | - |
|  | Observations | 4550 | 1782 | - |
| 2014 | Households | 2346 | 860 | - |
|  | Observations | 4099 | 1573 | - |
| 2015 | Households | 2408 | 889 | - |
|  | Observations | 4185 | 1620 | - |
| 2016 | Households | 2273 | 847 | - |
|  | Observations | 3940 | 1592 | - |
| 2017 | Households | 1950 | 619 | 58 |
|  | Observations | 3412 | 1188 | 102 |
| 2018 | Households | 1770 | 465 | 149 |
|  | Observations | 3086 | 899 | 282 |
| 2019 | Households | 1611 | 293 | 230 |
|  | Observations | 2830 | 570 | 424 |
| 2020 | Households | 1495 | 175 | 240 |
|  | Observations | 2618 | 346 | 452 |
| 2021 | Households | 1331 | 80 | 249 |
|  | Observations | 2271 | 141 | 461 |
| 2022 | Households | 1417 | 31 | 309 |
|  | Observations | 2401 | 55 | 569 |
| 2023 | Households | 1653 | - | 341 |
|  | Observations | 2754 | - | 651 |
| 2024 | Households | 655 | - | 170 |
|  | Observations | 1095 | - | 340 |
| 2025 | Households | 26 | - | 10 |
|  | Observations | 34 | - | 13 |

For 2017: cutoff based on interview date, as month of birth information not available. “Affected” corresponds to individuals whose 3^rd^ child was born after the cutoff (to our best approximation), and “unaffected” refers to those whose third or more child had been born approximately before.

#### Table S3: Raw means (90% CIs) of all outcomes by group by year

| Year |  | Mental health | | General health | | | Financial health | |
| --- | --- | --- | --- | --- | --- | --- | --- | --- |
|  |  | Psychological distress | Life satisfaction | Self-rated health | Health satisfaction | HRQoL | Financial situation | Financial outlook |
| 2009 | 1-2 children | 11.08 (10.91 - 11.25) | 2.79 (2.75 - 2.84) | 2.35 (2.32 - 2.38) | 2.90 (2.85 - 2.95) | 26.83 (26.51 - 27.16) | 2.55 (2.52 - 2.59) | 1.71 (1.69 - 1.73) |
|  | 3+ children | 11.38 (11.07 - 11.69) | 3.03 (2.95 - 3.11) | 2.49 (2.43 - 2.54) | 3.17 (3.07 - 3.26) | 28.38 (27.78 - 28.98) | 2.78 (2.72 - 2.83) | 1.78 (1.74 - 1.81) |
| 2010 | 1-2 children | 11.08 (10.96 - 11.20) | 2.82 (2.79 - 2.85) | 2.38 (2.35 - 2.40) | 3.00 (2.96 - 3.03) | 23.95 (23.74 - 24.16) | 2.49 (2.47 - 2.52) | 1.73 (1.72 - 1.74) |
|  | 3+ children | 11.35 (11.14 - 11.56) | 2.93 (2.87 - 2.98) | 2.46 (2.42 - 2.50) | 3.09 (3.03 - 3.16) | 25.34 (24.97 - 25.70) | 2.69 (2.65 - 2.72) | 1.77 (1.75 - 1.79) |
| 2011 | 1-2 children | 11.05 (10.93 - 11.18) | 2.82 (2.79 - 2.86) | 2.33 (2.31 - 2.35) | 3.20 (3.16 - 3.24) | 21.68 (21.50 - 21.85) | 2.49 (2.47 - 2.51) | 1.79 (1.77 - 1.80) |
|  | 3+ children | 11.42 (11.21 - 11.63) | 2.99 (2.94 - 3.05) | 2.43 (2.40 - 2.47) | 3.28 (3.22 - 3.35) | 22.90 (22.58 - 23.21) | 2.71 (2.67 - 2.75) | 1.83 (1.81 - 1.86) |
| 2012 | 1-2 children | 11.05 (10.92 - 11.18) | 2.88 (2.85 - 2.92) | 2.29 (2.27 - 2.32) | 3.34 (3.30 - 3.38) | 21.68 (21.51 - 21.86) | 2.47 (2.44 - 2.49) | 1.76 (1.74 - 1.77) |
|  | 3+ children | 11.48 (11.24 - 11.71) | 2.99 (2.93 - 3.05) | 2.43 (2.39 - 2.47) | 3.36 (3.29 - 3.43) | 22.78 (22.46 - 23.09) | 2.67 (2.63 - 2.71) | 1.82 (1.80 - 1.85) |
| 2013 | 1-2 children | 11.09 (10.95 - 11.22) | 2.89 (2.85 - 2.92) | 2.33 (2.30 - 2.35) | 3.33 (3.29 - 3.38) | 21.74 (21.57 - 21.91) | 2.42 (2.40 - 2.45) | 1.75 (1.74 - 1.77) |
|  | 3+ children | 11.75 (11.49 - 12.00) | 3.01 (2.95 - 3.08) | 2.42 (2.38 - 2.46) | 3.36 (3.28 - 3.43) | 22.90 (22.58 - 23.23) | 2.63 (2.58 - 2.67) | 1.81 (1.79 - 1.84) |
| 2014 | 1-2 children | 10.93 (10.79 - 11.08) | 2.80 (2.76 - 2.84) | 2.32 (2.29 - 2.34) | 3.23 (3.19 - 3.28) | 21.52 (21.34 - 21.70) | 2.31 (2.29 - 2.34) | 1.71 (1.70 - 1.73) |
|  | 3+ children | 11.45 (11.19 - 11.71) | 2.94 (2.88 - 3.01) | 2.41 (2.36 - 2.45) | 3.35 (3.27 - 3.42) | 22.72 (22.36 - 23.07) | 2.50 (2.46 - 2.54) | 1.75 (1.73 - 1.78) |
| 2015 | 1-2 children | 10.73 (10.59 - 10.87) | 2.68 (2.65 - 2.72) | 2.31 (2.29 - 2.34) | 2.97 (2.93 - 3.01) | 21.84 (21.64 - 22.03) | 2.21 (2.18 - 2.23) | 1.68 (1.67 - 1.70) |
|  | 3+ children | 11.19 (10.95 - 11.43) | 2.85 (2.78 - 2.91) | 2.41 (2.37 - 2.45) | 3.06 (2.99 - 3.14) | 23.32 (22.96 - 23.68) | 2.41 (2.37 - 2.45) | 1.74 (1.72 - 1.77) |
| 2016 | 1-2 children | 11.02 (10.87 - 11.16) | 2.73 (2.70 - 2.77) | 2.41 (2.39 - 2.44) | 2.92 (2.88 - 2.96) | 21.88 (21.68 - 22.07) | 2.18 (2.15 - 2.20) | 1.70 (1.68 - 1.72) |
|  | 3+ children | 11.21 (10.97 - 11.46) | 2.81 (2.75 - 2.88) | 2.46 (2.42 - 2.50) | 2.97 (2.90 - 3.04) | 23.30 (22.95 - 23.66) | 2.32 (2.28 - 2.36) | 1.75 (1.72 - 1.77) |
| 2017 | 1-2 children | 11.35 (11.19 - 11.51) | 2.80 (2.77 - 2.84) | 2.45 (2.43 - 2.48) | 3.03 (2.99 - 3.08) | 22.15 (21.94 - 22.37) | 2.24 (2.21 - 2.26) | 1.70 (1.68 - 1.72) |
|  | 3+ children | 11.23 (10.72 - 11.75) | 2.87 (2.73 - 3.01) | 2.48 (2.39 - 2.57) | 3.02 (2.86 - 3.17) | 23.16 (22.43 - 23.90) | 2.31 (2.22 - 2.40) | 1.73 (1.67 - 1.78) |
|  | 3+ children (affected) | 11.30 (10.26 - 12.34) | 2.89 (2.65 - 3.13) | 2.43 (2.26 - 2.60) | 3.02 (2.76 - 3.28) | 23.01 (21.58 - 24.44) | 2.82 (2.63 - 3.01) | 1.80 (1.69 - 1.91) |
|  | 3+ children (unaffected) | 11.04 (10.73 - 11.36) | 2.90 (2.82 - 2.99) | 2.58 (2.52 - 2.64) | 3.08 (2.98 - 3.18) | 22.94 (22.48 - 23.40) | 2.47 (2.41 - 2.52) | 1.78 (1.74 - 1.81) |
| 2018 | 1-2 children | 11.56 (11.39 - 11.73) | 2.83 (2.79 - 2.87) | 2.52 (2.49 - 2.55) | 3.06 (3.01 - 3.10) | 22.39 (22.16 - 22.61) | 2.26 (2.23 - 2.29) | 1.73 (1.72 - 1.75) |
|  | 3+ children (affected) | 11.35 (10.73 - 11.98) | 2.95 (2.79 - 3.11) | 2.49 (2.38 - 2.59) | 2.99 (2.83 - 3.16) | 23.47 (22.64 - 24.30) | 2.69 (2.59 - 2.79) | 1.76 (1.70 - 1.82) |
|  | 3+ children (unaffected) | 11.84 (11.49 - 12.19) | 3.07 (2.98 - 3.15) | 2.62 (2.56 - 2.67) | 3.25 (3.15 - 3.35) | 23.64 (23.18 - 24.10) | 2.43 (2.38 - 2.49) | 1.79 (1.75 - 1.82) |
| 2019 | 1-2 children | 11.72 (11.54 - 11.90) | 2.82 (2.78 - 2.86) | 2.52 (2.49 - 2.55) | 3.06 (3.01 - 3.11) | 23.00 (22.76 - 23.23) | 2.25 (2.22 - 2.28) | 1.75 (1.73 - 1.77) |
|  | 3+ children (affected) | 11.64 (11.14 - 12.14) | 3.01 (2.88 - 3.14) | 2.64 (2.56 - 2.72) | 3.18 (3.05 - 3.32) | 24.41 (23.71 - 25.11) | 2.58 (2.49 - 2.66) | 1.77 (1.72 - 1.82) |
|  | 3+ children (unaffected) | 12.09 (11.65 - 12.53) | 3.16 (3.05 - 3.26) | 2.69 (2.62 - 2.76) | 3.24 (3.12 - 3.36) | 24.63 (24.04 - 25.22) | 2.57 (2.50 - 2.65) | 1.84 (1.79 - 1.88) |
| 2020 | 1-2 children | 12.13 (11.95 - 12.32) | 2.75 (2.70 - 2.79) | 2.52 (2.49 - 2.55) | 3.03 (2.98 - 3.07) | 22.98 (22.75 - 23.22) | 2.16 (2.13 - 2.19) | 1.79 (1.77 - 1.81) |
|  | 3+ children (affected) | 12.74 (12.24 - 13.24) | 2.99 (2.87 - 3.10) | 2.67 (2.60 - 2.75) | 3.21 (3.08 - 3.33) | 24.38 (23.73 - 25.03) | 2.41 (2.33 - 2.49) | 1.83 (1.78 - 1.88) |
|  | 3+ children (unaffected) | 12.69 (12.10 - 13.28) | 2.99 (2.87 - 3.12) | 2.60 (2.51 - 2.69) | 3.05 (2.91 - 3.19) | 24.06 (23.30 - 24.83) | 2.39 (2.31 - 2.48) | 1.84 (1.78 - 1.89) |
| 2021 | 1-2 children | 11.99 (11.79 - 12.19) | 2.73 (2.69 - 2.78) | 2.52 (2.49 - 2.55) | 3.04 (2.99 - 3.09) | 22.79 (22.54 - 23.05) | 2.14 (2.11 - 2.17) | 1.75 (1.73 - 1.78) |
|  | 3+ children (affected) | 11.67 (11.22 - 12.11) | 2.91 (2.80 - 3.01) | 2.62 (2.55 - 2.70) | 3.15 (3.02 - 3.27) | 23.44 (22.84 - 24.04) | 2.33 (2.26 - 2.41) | 1.81 (1.76 - 1.86) |
|  | 3+ children (unaffected) | 11.71 (10.80 - 12.62) | 2.72 (2.53 - 2.91) | 2.55 (2.42 - 2.68) | 3.03 (2.81 - 3.25) | 22.72 (21.57 - 23.88) | 2.43 (2.29 - 2.56) | 1.70 (1.61 - 1.80) |
| 2022 | 1-2 children | 12.16 (11.96 - 12.36) | 2.82 (2.77 - 2.86) | 2.60 (2.56 - 2.63) | 3.09 (3.04 - 3.14) | 23.40 (23.14 - 23.67) | 2.33 (2.30 - 2.36) | 1.99 (1.96 - 2.01) |
|  | 3+ children (affected) | 12.20 (11.75 - 12.65) | 3.02 (2.92 - 3.12) | 2.66 (2.59 - 2.73) | 3.15 (3.04 - 3.26) | 24.13 (23.55 - 24.71) | 2.60 (2.53 - 2.68) | 2.03 (1.98 - 2.08) |
|  | 3+ children (unaffected) | 11.37 (10.00 - 12.75) | 2.82 (2.52 - 3.12) | 2.56 (2.38 - 2.74) | 2.57 (2.27 - 2.88) | 21.47 (20.10 - 22.84) | 2.66 (2.40 - 2.91) | 2.19 (2.01 - 2.36) |
| 2023 | 1-2 children | 12.19 (12.01 - 12.38) | 2.82 (2.78 - 2.86) | 2.59 (2.56 - 2.62) | 3.16 (3.11 - 3.21) | 23.63 (23.38 - 23.88) | 2.39 (2.36 - 2.42) | 1.86 (1.84 - 1.89) |
|  | 3+ children (affected) | 12.50 (12.11 - 12.90) | 3.15 (3.05 - 3.24) | 2.68 (2.62 - 2.74) | 3.24 (3.13 - 3.34) | 24.88 (24.33 - 25.44) | 2.63 (2.56 - 2.70) | 1.98 (1.93 - 2.02) |
| 2024 | 1-2 children | 12.51 (12.21 - 12.81) | 2.84 (2.77 - 2.90) | 2.66 (2.61 - 2.71) | 3.17 (3.10 - 3.25) | 24.44 (24.03 - 24.84) | 2.37 (2.33 - 2.42) | 1.80 (1.76 - 1.83) |
|  | 3+ children (affected) | 12.51 (11.92 - 13.10) | 3.17 (3.04 - 3.31) | 2.78 (2.70 - 2.86) | 3.36 (3.21 - 3.50) | 25.27 (24.51 - 26.02) | 2.65 (2.55 - 2.74) | 1.89 (1.83 - 1.95) |
| 2025 | 1-2 children | 12.41 (11.09 - 13.72) | 2.97 (2.56 - 3.37) | 2.79 (2.46 - 3.12) | 3.17 (2.71 - 3.63) | 24.19 (21.43 - 26.96) | 2.29 (1.94 - 2.65) | 1.71 (1.54 - 1.87) |
|  | 3+ children (affected) | 10.15 (8.15 - 12.16) | 2.92 (2.27 - 3.58) | 2.54 (2.18 - 2.89) | 2.92 (2.32 - 3.53) | 24.00 (20.30 - 27.70) | 2.17 (1.74 - 2.61) | 1.74 (1.45 - 2.03) |

For 2017: cutoff based on interview date, as month of birth information not available. “Affected” corresponds to individuals whose 3^rd^ child was born after the cutoff (to our best approximation), and “unaffected” refers to those whose third child had been born approximately before.

#### Table S4: ITS (change in slope per year) results of all outcomes (90% CIs)

|  | Unadjusted | Adjusted | Standardised adjusted |
| --- | --- | --- | --- |
|  | Change in slope / year (90% CI) | Change in slope / year (90% CI) | Change in slope / year (90% CI) |
| Psychological distress | 0.09 (-0.07, 0.25) | 0.09 (-0.06, 0.24) | 0.02 (-0.01, 0.04) |
| Life satisfaction | 0.02 (-0.02, 0.07) | 0.02 (-0.02, 0.06) | 0.01 (-0.01, 0.04) |
| Self-rated health | 0.01 (-0.02, 0.04) | 0.01 (-0.02, 0.03) | 0.01 (-0.02, 0.03) |
| HRQoL | 0.67 (0.43, 0.92) | 0.65 (0.41, 0.88) | 0.08 (0.05, 0.11) |
| Health satisfaction | 0.00 (-0.04, 0.05) | 0.00 (-0.04, 0.04) | 0.00 (-0.02, 0.03) |
| Financial situation | 0.08 (0.05, 0.11) | 0.08 (0.05, 0.11) | 0.08 (0.05, 0.11) |
| Financial outlook | 0.03 (0.01, 0.05) | 0.03 (0.01, 0.05) | 0.04 (0.01, 0.07) |

#### Table S5: Parallel trends numerical results (90% CIs) for all outcomes

|  | Unadjusted | Adjusted | Standardised adjusted |
| --- | --- | --- | --- |
|  | Family size group indicator*year (90% CI) | Family size group indicator*year (90% CI) | Family size group indicator*year (90% CI) |
| Psychological distress | 0.05 (-0.02, 0.12) | 0.05 (-0.02, 0.12) | 0.01 (-0.00, 0.02) |
| Life satisfaction | 0.00 (-0.02, 0.02) | -0.00 (-0.02, 0.02) | -0.00 (-0.01, 0.01) |
| Self-rated health | 0.00 (-0.01, 0.02) | -0.00 (-0.01, 0.01) | -0.00 (-0.01, 0.01) |
| HRQoL | 0.05 (-0.08, 0.17) | 0.03 (-0.09, 0.14) | 0.00 (-0.01, 0.02) |
| Health satisfaction | -0.01 (-0.04, 0.01) | -0.01 (-0.03, 0.01) | -0.01 (-0.02, 0.01) |
| Financial situation | 0.01 (-0.01, 0.02) | -0.00 (-0.02, 0.01) | -0.00 (-0.02, 0.01) |
| Financial outlook | -0.00 (-0.01, 0.01) | 0.00 (-0.01, 0.01) | 0.00 (-0.01, 0.01) |

#### Table S6: Parallel trends numerical results (90% CIs) for subgroups in all standardised adjusted outcomes

|  | Household income | | Sex | | Ethnicity | |
| --- | --- | --- | --- | --- | --- | --- |
|  | Income-low | Income-high | Male | Female | White | Ethnic Minority |
|  | Family size group indicator*year (90% CI) | Family size group indicator*year (90% CI) | Family size group indicator*year (90% CI) | Family size group indicator*year (90% CI) | Family size group indicator*year (90% CI) | Family size group indicator*year (90% CI) |
| Psychological distress | 0.00 (-0.01, 0.02) | 0.02 (0.00, 0.03) | 0.01 (-0.01, 0.03) | 0.01 (-0.01, 0.02) | 0.01 (-0.01, 0.02) | 0.02 (-0.00, 0.05) |
| Life satisfaction | -0.01 (-0.03, 0.01) | 0.01 (-0.00, 0.03) | -0.00 (-0.02, 0.02) | 0.00 (-0.02, 0.02) | 0.00 (-0.01, 0.02) | 0.00 (-0.03, 0.03) |
| Self-rated health | 0.00 (-0.02, 0.02) | 0.01 (-0.01, 0.03) | -0.01 (-0.02, 0.02) | 0.00 (-0.01, 0.02) | 0.00 (-0.01, 0.02) | -0.01 (-0.03, 0.01) |
| HRQoL | 0.01 (-0.01, 0.03) | 0.01 (-0.01, 0.03) | -0.01 (-0.03, 0.02) | 0.01 (-0.01, 0.03) | -0.00 (-0.02, 0.02) | 0.02 (-0.01, 0.05) |
| Health satisfaction | -0.02 (-0.04, 0.00) | 0.01 (-0.01, 0.02) | -0.02 (-0.04, -0.00) | 0.01 (-0.01, 0.02) | -0.00 (-0.02, 0.01) | -0.03 (-0.05, -0.00) |
| Financial situation | -0.01 (-0.03, 0.02) | 0.01 (-0.01, 0.03) | 0.01 (-0.01, 0.03) | -0.01 (-0.02, 0.01) | -0.01 (-0.02, 0.01) | 0.03 (-0.00, 0.06) |
| Financial outlook | -0.00 (-0.02, 0.02) | -0.00 (-0.02, 0.02) | -0.01 (-0.03, 0.01) | 0.01 (-0.00, 0.02) | 0.00 (-0.01, 0.02) | -0.01 (-0.03, 0.02) |

#### Table S7: DiD (difference in change in slope) results (90% CIs) for all outcomes

|  | Parallel trends assumption met? (for standardised adjusted model) | Unadjusted | Adjusted | Standardised adjusted |
| --- | --- | --- | --- | --- |
|  |  | Family size group indicator*pre-post intervention time indicator (90% CI) | Family size group indicator *pre-post intervention time indicator (90% CI) | Family size group indicator *pre-post intervention time indicator (90% CI) |
| Psychological distress | Y | -0.24 (-1.05, 0.58) | -0.13 (-0.92, 0.67) | -0.02 (-0.16, 0.12) |
| Life satisfaction | Y | 0.16 (-0.04, 0.37) | 0.17 (-0.03, 0.38) | 0.12 (-0.02, 0.27) |
| Self-rated health | Y | 0.01 (-0.11, 0.13) | 0.02 (-0.10, 0.14) | 0.02 (-0.01, 0.14) |
| HRQoL | Y | -0.07 (-1.24, 1.10) | 0.00 (-1.15, 1.16) | 0.00 (-0.14, 0.14) |
| Health satisfaction | Y | -0.06 (-0.25, 0.14) | -0.04 (-0.23, 0.15) | -0.03 (-0.14, 0.09) |
| Financial situation | Y | 0.08 (-0.08, 0.25) | 0.09 (-0.06, 0.24) | 0.09 (-0.06, 0.24) |
| Financial outlook | Y | 0.04 (-0.07, 0.14) | 0.04 (-0.06, 0.14) | 0.06 (-0.09, 0.21) |

#### Table S8: CITS (difference in change of slope per year) results (90% CIs) for all outcomes

|  | Unadjusted | Adjusted | Standardised-adjusted |
| --- | --- | --- | --- |
|  | Difference in change in slope of 3+ children group and change in slope of 1-2 children group / year (90% CI) | Difference in change in slope of 3+ children group and change in slope of 1-2 children group / year (90% CI) | Difference in change in slope of 3+ children group and change in slope of 1-2 children group / year (90% CI) |
| Psychological distress | -0.01 (-0.17, 0.14) | -0.00 (-0.15, 0.15) | -0.00 (-0.03, 0.03) |
| Life satisfaction | 0.00 (-0.04, 0.04) | 0.01 (-0.03, 0.05) | 0.01 (-0.02, 0.04) |
| Self-rated health | -0.01 (-0.04, 0.02) | -0.01 (-0.03, 0.02) | -0.01 (-0.03, 0.02) |
| HRQoL | -0.04 (-0.28, 0.20) | 0.02 (-0.22, 0.25) | 0.00 (-0.03, 0.03) |
| Health satisfaction | -0.01 (-0.06, 0.03) | -0.01 (-0.05, 0.03) | -0.01 (-0.03, 0.02) |
| Financial situation | -0.00 (-0.03, 0.03) | 0.01 (-0.02, 0.04) | 0.01 (-0.02, 0.04) |
| Financial outlook | -0.00 (-0.02, 0.02) | -0.01 (-0.02, 0.01) | -0.01 (-0.04, 0.02) |

#### Table S9: Sensitivity analyses without weights applied for all standardised adjusted outcomes (90% CIs)

|  | ITS | DiD | CITS |
| --- | --- | --- | --- |
|  | Change in slope / year (90% CI) | Family size group indicator *pre-post intervention time indicator (90% CI) | Difference in change in slope of 3+ children group and change in slope of 1-2 children group / year (90% CI) |
| Psychological distress | 0.04 (0.01, 0.06) | 0.01 (-0.11, 0.12) | 0.01 (-0.01, 0.03) |
| Life satisfaction | 0.03 (0.01, 0.05) | 0.13 (0.02, 0.24) | 0.02 (0.00, 0.05) |
| Self-rated health | 0.02 (0.00, 0.04) | 0.04 (-0.06, 0.13) | 0.01 (-0.01, 0.03) |
| HRQoL | 0.09 (0.06, 0.11) | -0.02 (-0.14, 0.09) | 0.01 (-0.02, 0.03) |
| Health satisfaction | 0.02 (-0.00, 0.04) | 0.02 (-0.07, 0.12) | 0.01 (-0.01, 0.03) |
| Financial situation | 0.09 (0.07, 0.12) | 0.07 (-0.05, 0.19) | 0.02 (-0.00, 0.05) |
| Financial outlook | 0.06 (0.03, 0.08) | 0.07 (-0.04, 0.18) | -0.00 (-0.02, 0.02) |

#### Table S10: DiD falsification test standardised analyses (90% CIs)

|  | Family size group indicator *pre-post intervention time indicator (90% CI) |
| --- | --- |
| Psychological distress | 0.01 (-0.18, 0.20) |
| Life satisfaction | -0.08 (-0.30, 0.14) |
| Self-rated health | -0.13 (-0.32, 0.06) |
| HRQoL | -0.05 (-0.22, 0.13) |
| Health satisfaction | -0.06 (-0.26, 0.13) |
| Financial situation | -0.04 (-0.29, 0.21) |
| Financial outlook | -0.06 (-0.26, 0.14) |

#### Table S11: DiD + PSM standardised adjusted analyses (90% CIs)

|  | Family size group indicator *pre-post intervention time indicator (90% CI) |
| --- | --- |
| Psychological distress | -0.04 (-0.13, 0.05) |
| Life satisfaction | 0.07 (-0.01, 0.15) |
| Self-rated health | 0.02 (-0.07, 0.10) |
| HRQoL | -0.02 (-0.10, 0.06) |
| Health satisfaction | 0.01 (-0.07, 0.08) |
| Financial situation | 0.02 (-0.07, 0.10) |
| Financial outlook | 0.02 (-0.06, 0.10) |

### Supplementary Figures

#### Figure S1: Sample selection chart

601,795 total observations in adult sample

564,989 observations with fully or partially-productive interviews

564,987 observations between 2009 and 2025 (inclusive)

78,015 observations with $\geq$ 1 child 5 or under

77,877 observations with at least one outcome response

76,878 observations whose 3rd or more child was not born in 2017

#### Figure S2: Outcome correlation diagram

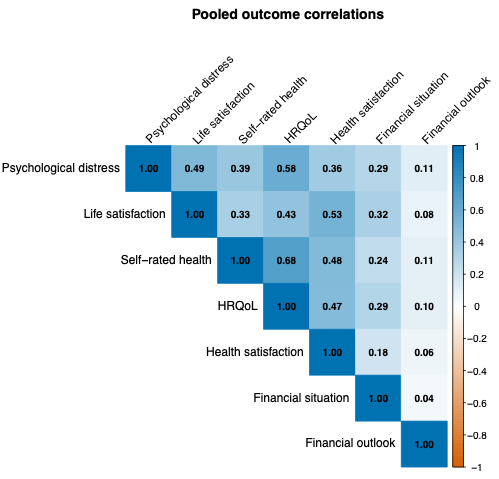

#### Figure S3: Stacked bar graphs showing proportion of observations with 3+ children affected by the two-child benefit cap every year from 2017

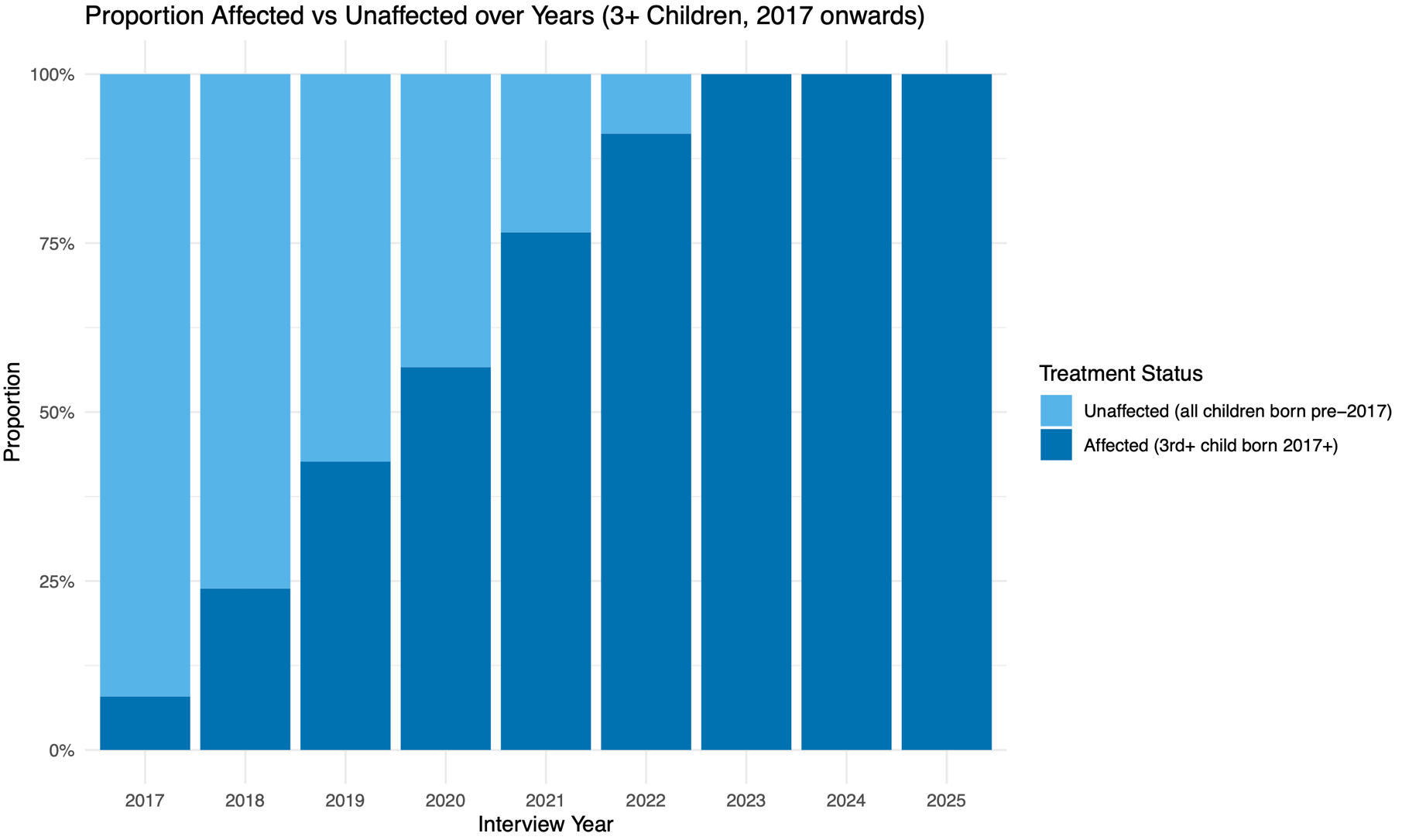

Note that this figure is based on birth year, as month and day of birth information was not available.

#### Figure S4: Interrupted time series results across all outcomes with 3+ children group (raw adjusted)

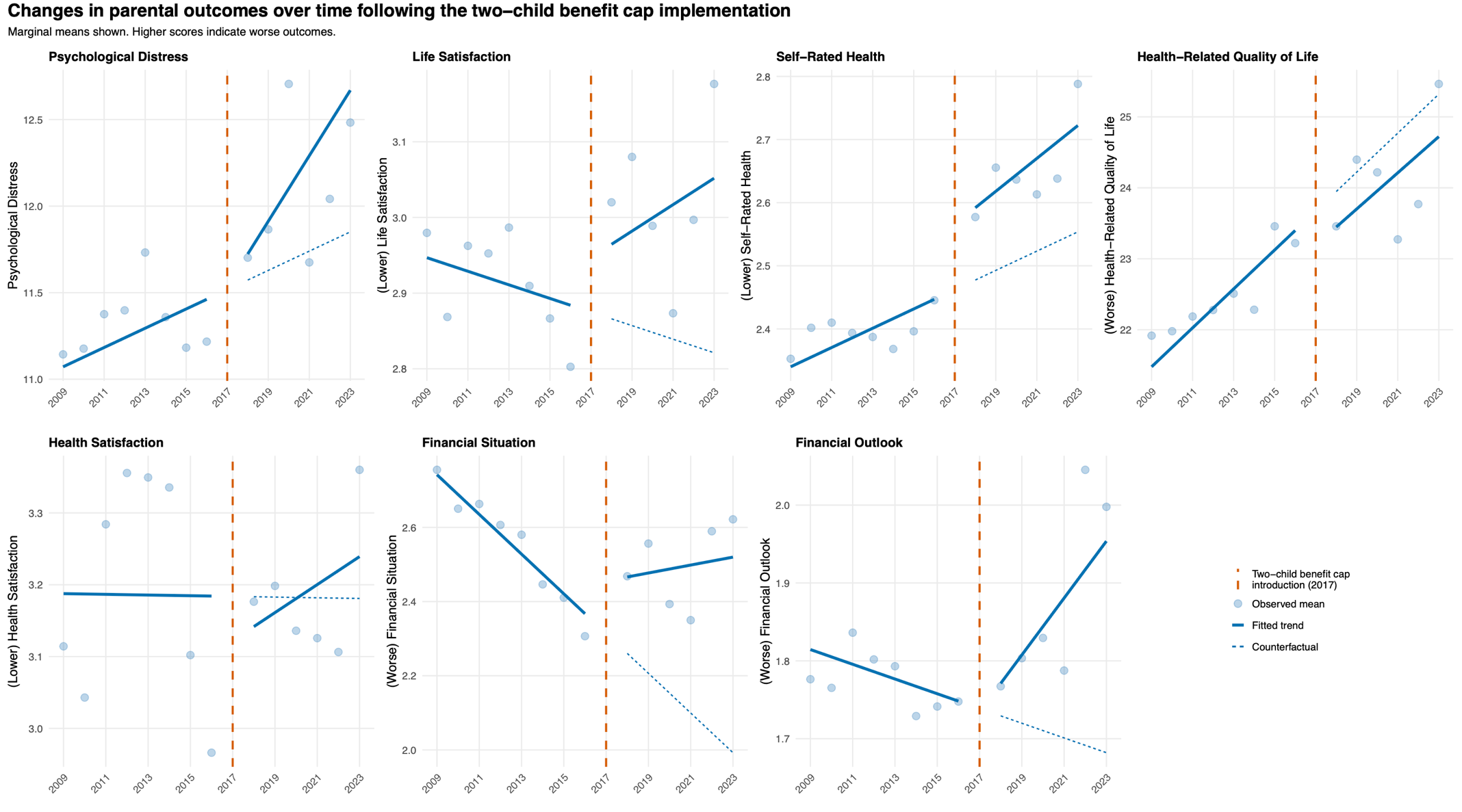

#### Figure S5: Visualisation of standardised parallel trends between 2009 and 2016 per year for 3+ children group compared to 1-2 children group

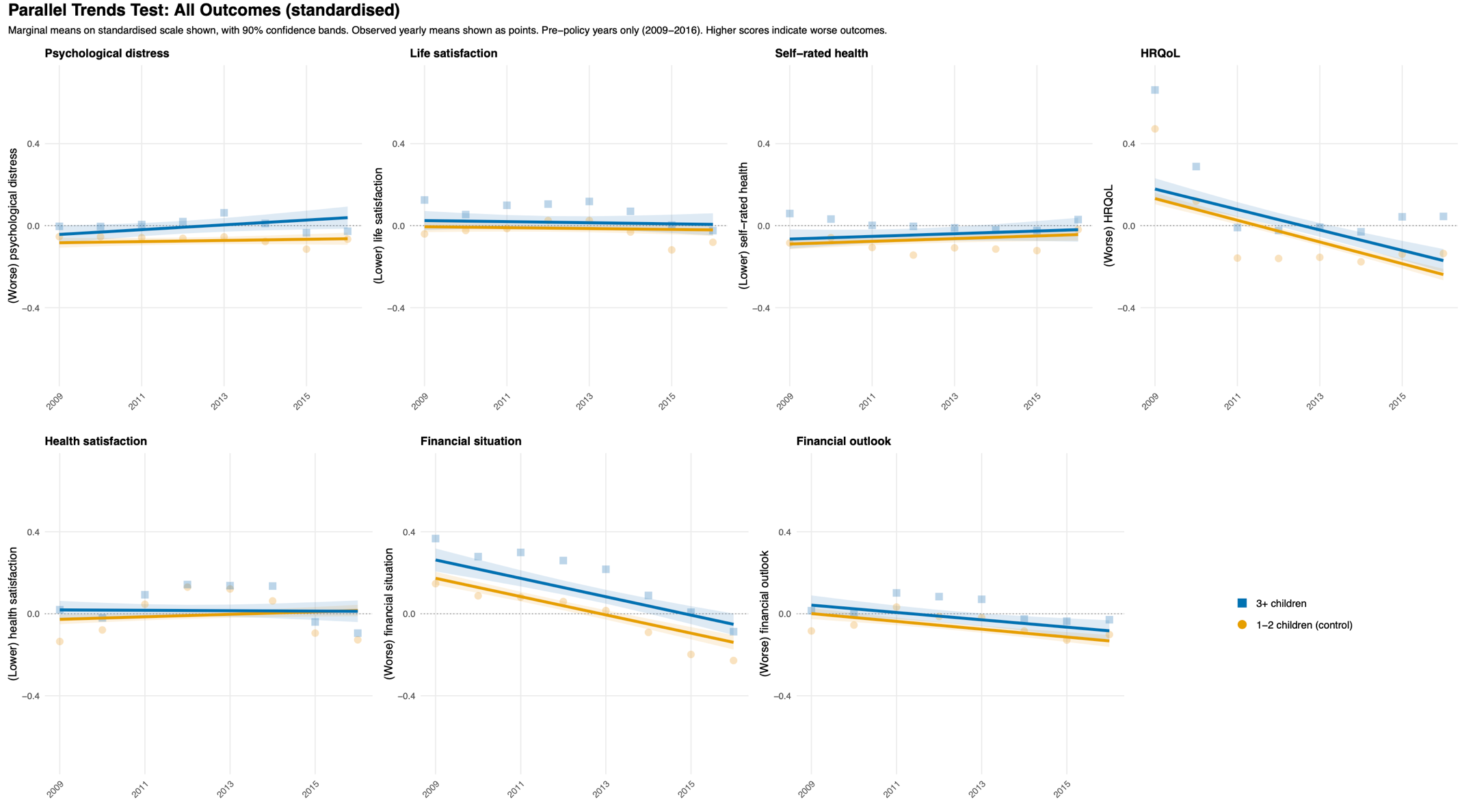

#### Figure S6: Mean total scores for difference-in-differences analyses between 2014-2016 and 2023-2025 (raw adjusted)

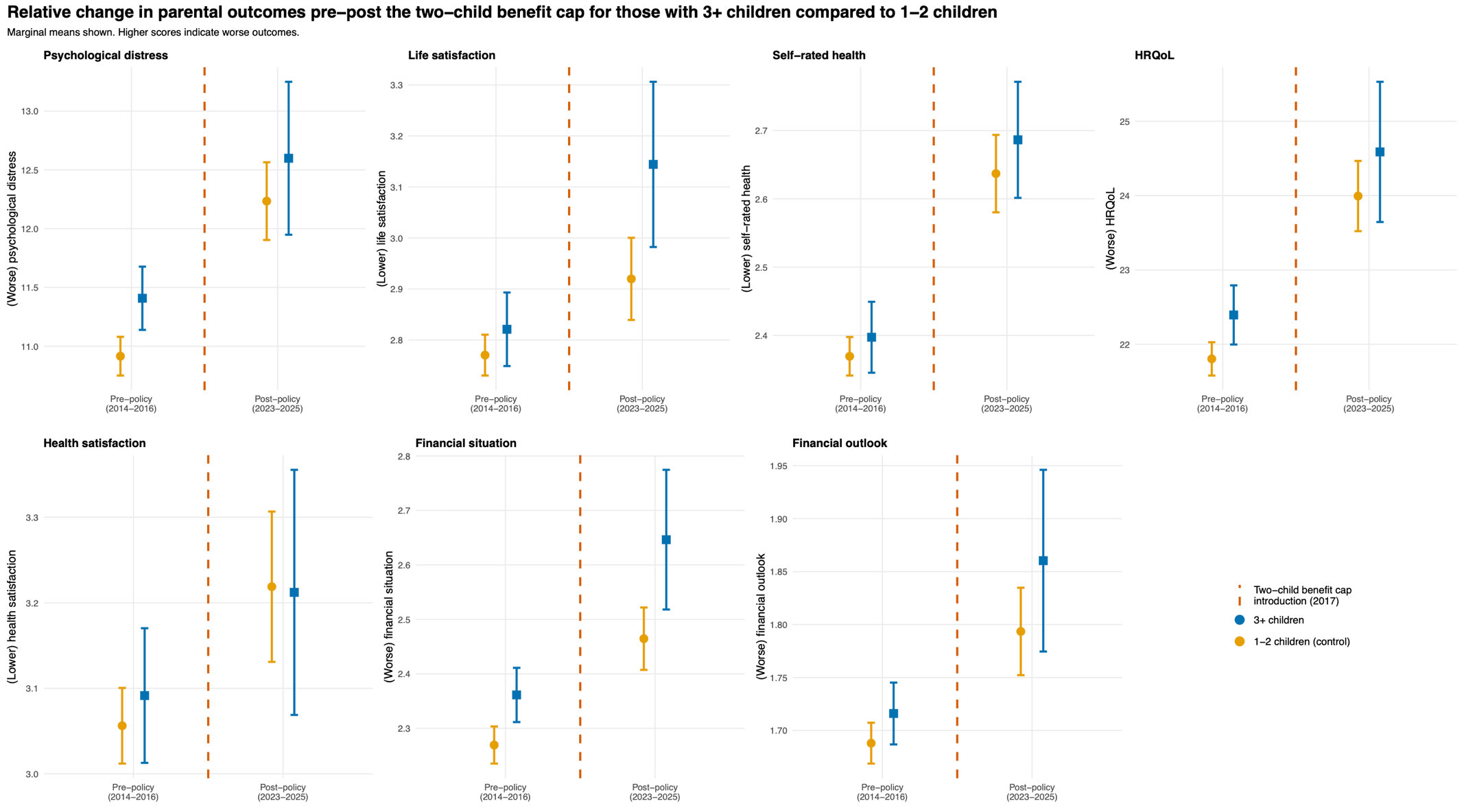

#### Figure S7: Controlled interrupted time series results across all outcomes with 3+ children compared to 1-2 children group (raw adjusted)

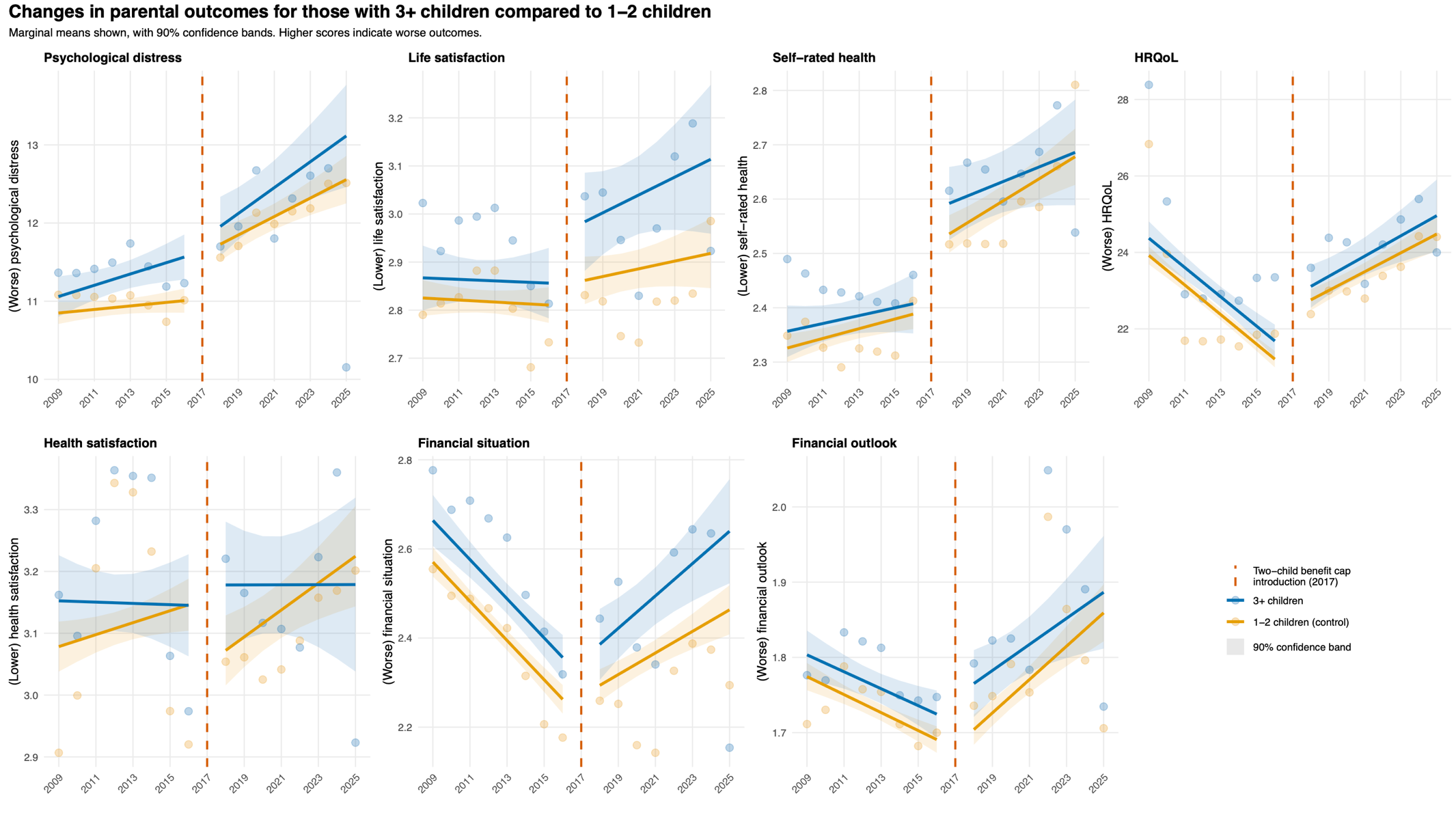

#### Figure S8: Mean total scores for propensity-score-matched difference-in-differences analyses between 2013-2016 and 2018-2025 (standardised adjusted)

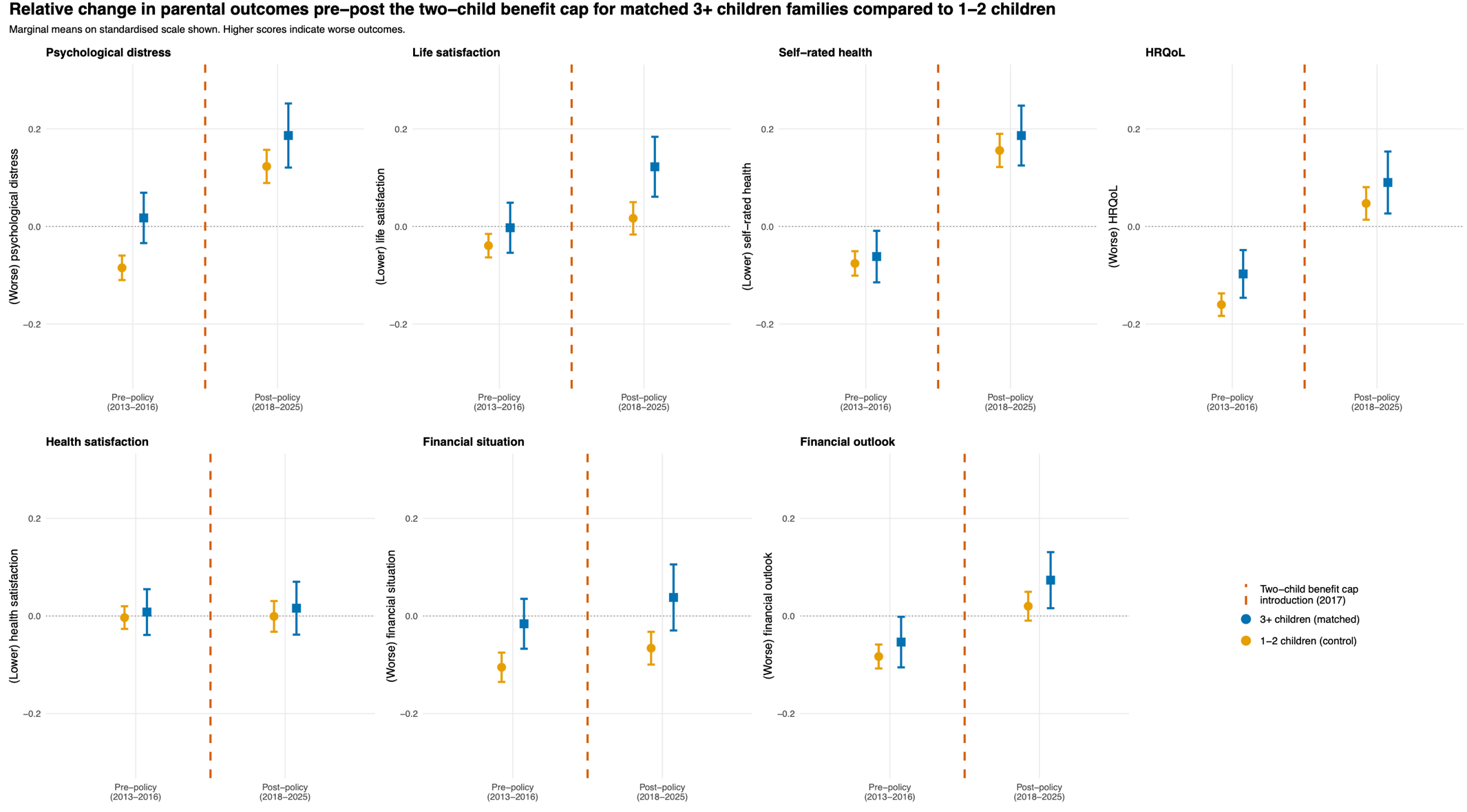

### Supplementary References

1.	University of Essex. Survey timeline. *Underst Soc* n.d. https://www.understandingsociety.ac.uk/documentation/mainstage/survey-timeline/ (28 Aug. 2026, date last accessed).
